# Seasonal malaria chemoprevention and the spread of *Plasmodium falciparum* quintuple mutant parasites resistant to sulfadoxine-pyrimethamine: a modelling study

**DOI:** 10.1101/2023.07.23.23293041

**Authors:** Thiery Masserey, Tamsin Lee, Sherrie L Kelly, Ian M Hastings, Melissa A Penny

**Affiliations:** Swiss Tropical and Public Health Institute, Allschwil, Switzerland; University of Basel, Basel, Switzerland; Liverpool School of Tropical Medicine, Liverpool, UK

## Abstract

**Background:** Seasonal malaria chemoprevention (SMC) with sulfadoxine-pyrimethamine (SP) plus amodiaquine (AQ) prevents millions of clinical malaria cases in children under five in Africa’s Sahel region. However, parasites partially resistant to SP (with “quintuple” mutations) potentially threaten SMC protective effectiveness. We evaluated its spread and clinical consequences.

**Methods:** An individual-based malaria transmission model with explicit parasite dynamics and drug pharmacological models, was used to identify and quantify the influence of factors driving quintuple mutant spread and predict the time needed for the mutant to spread from 1% to 50% of inoculations for several SMC deployment strategies. We estimated the impact of this spread on SMC effectiveness against clinical malaria.

**Findings:** Higher transmission intensity, SMC coverage, and expanded age range of chemoprevention promoted mutant spread. SMC implementation in a high transmission setting (40% parasite prevalence in children aged 2-10 years) with four monthly cycles to children aged three months to five years (with 95% initial coverage declining each cycle), the mutant requires 53·1 years (95% CI 50·5–56·0) to spread from 1% to 50% of inoculations. This time increased in lower transmission settings and reduced by half when SMC was extended to children under ten, or reduced by 10-13 years when an additional monthly cycle of SMC was deployed. For the same setting, the effective reduction in clinical cases in children receiving SMC was 79·0% (95% CI 77·8–80·8) and 60·4% (95% CI 58·6–62·3) during the months of SMC implementation when the mutant was absent or fixed in the population, respectively.

**Interpretation:** SMC with SP+AQ leads to a relatively slow spread of SP-resistant quintuple mutants and remains effective at preventing clinical malaria despite the mutant spread. SMC with SP+AQ should be considered in seasonal settings where this mutant is already prevalent.

**Funding:** Swiss National Science Foundation and Marie Curie Individual Fellowship.

## Introduction

*Plasmodium falciparum* malaria is a leading cause of morbidity and mortality in African children.^1^ In Africa’s Sahel region, *Plasmodium falciparum* transmission is highly seasonal, with most clinical cases occurring over a three to five month period.^2^ Since 2012, the World Health Organization (WHO) has recommended implementation of seasonal malaria chemoprevention (SMC) in this region,^2^ and recent updated guidelines are paving the way for more flexible SMC implementation (varying the number of cycles and targeted age groups)^3^ SMC has been implemented as monthly sulfadoxine and pyrimethamine (SP) plus amodiaquine (AQ) to children aged three months to five years during the transmission season.^2^ A large implementation study in the Sahel reported that SMC prevented over 88% of uncomplicated malaria cases within 28-days of administration.^4^ This high effectiveness is partly attributable to the fact that SP remains at a concentration sufficient to inhibit development of successful blood-stage infections for long periods post-treatment. Current evidence suggests that this prophylactic period (PP) is roughly 42-days against SP-sensitive parasites, but shorter for less sensitive parasites.^5–7^

Accumulation of mutations in *dihydropteroate synthase (dhps)* and *dihydrofolate reductase (dhfr) P. falciparum* genes leads to reduced sensitivity to sulfadoxine and pyrimethamine, respectively.^8^ In many Sahelian countries, the quadruple mutant (*dhfr*-51I, *dhfr*-59A, *dhfr*- 108A, and *dhps-*437G) is already highly prevalent.^4, 9, 10^ It is challenging to estimate the exact PP conferred by SP against a specific mutant in the real world due to the presence of other mutants, geographical variation in mutant frequency, and individual variation. However, in West Africa, where the quadruple mutant is highly prevalent, SP still provides an important protection period of approximately 35-days (figure 1A) in clinical trials of SMC with SP+AQ^11^ and in a prospective study of intermittent preventive treatment in pregnancy (IPTp) with SP.^5^

**Figure 1:**
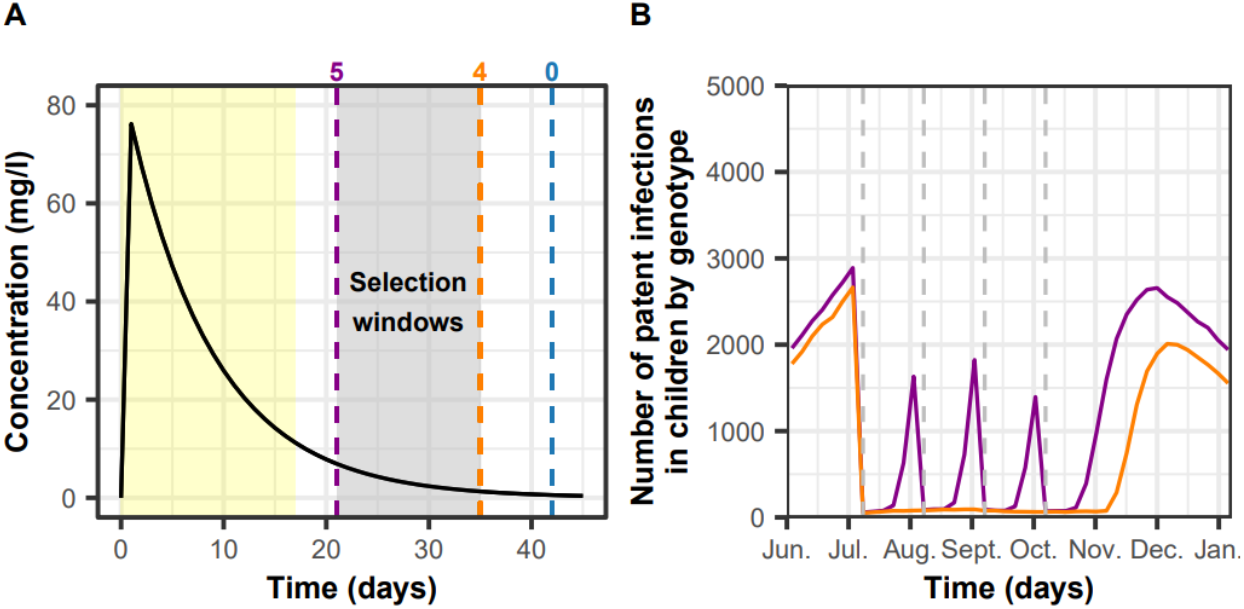
Illustrations of SP resistance calibration and dynamics modelling. (A) Within-host plasma concentration of SP* (the single long-acting drug with a similar duration of action as the expected synergetic combination of sulfadoxine and pyrimethamine) was modelled as a single long-acting drug with a one-compartment PK/PD model with first-order absorption (appendix p 8-11 table S1.3). SP* mimicked the synergic effect of SP combinations on the blood-stage (appendix p 8-11 table S1.3). The purple, orange, and blue dashed lines represent the end of the PP for a quintuple mutant, quadruple mutant, and a sensitive parasite, respectively (obtained from empirical studies).^5–7, 11^ EC50 values of 24·0 mg/l for the quintuple mutant and 2·4 mg/l for the quadruple mutant confer these clinically-observed PPs (appendix p8-11). The yellow region represents the PP conferred by AQ and its active metabolite against all genotypes.^12^ AQ and its active metabolite were modelled separately using two two-compartment models with first-order absorption (appendix p 12-16 tables S1.4 and S1.5). The grey region between the orange and purple lines highlights the selection window caused by SP (period during which the quintuple mutant can develop a successful blood-stage infection in SMC-treated children, but the quadruple mutant cannot). (B) Example of the predicted number of patent infections (defined as infections detectable by microscopy) in children aged three months to five years in a population consisting of 50% of quadruple mutant (orange line) and 50% of quintuple (purple line) mutant in a setting with a transmission intensity of 390 inoculations per person per year occurring mainly over four months (representing the seasonality pattern of Burkina Faso) and with a 35% probability of symptomatic cases receiving treatment within two weeks from symptom onset (defined as the level of access to treatment). Grey dashed lines indicate timing for cycles of SMC administered. In this example, four cycles of SMC were administered monthly with a coverage of 98% to children aged three months to ten years with no reduction of coverage between cycles.

A bigger threat comes from emergence of a quintuple mutant in multiple countries of the Sahel.^4^ This mutant carries an additional mutation (*dhps-*540G) conferring higher resistance to SP leading to high treatment failure rates with the use of SP monotherapy.^8^ However, clinical trial data of intermittent preventive treatment in infancy (IPTi) with SP^6, 8^ and a prospective study of IPTp with SP^5^ suggest that SP prevents successful development of a quintuple mutant blood-stage infection (due to reinfection or recrudescence) for 21-days post-treatment (figure 1A). Quintuple mutants can establish blood-stage infections more rapidly post-treatment than more sensitive parasites, so implementation of SMC may drive its spread (figure 1A). This selection occurs even when SP is used in combination with AQ because AQ provides an approximately 17-day PP; therefore, AQ is eliminated before the selection window caused by SP (period during which the quintuple mutant can develop a successful blood-stage infection in SMC-treated children, but the quadruple mutant cannot) occurs (figure 1A).^12^ Note that markers of low degrees of resistance to AQ (*Pfcrt-CIVET* + *pfmdr1-86 Tyr* + *184 Tyr*), which can slightly reduce AQ’s prophylactic action, have been observed in the Sahel at a low prevalence (0·5% in 2018).^4, 12^ However, this is unlikely to change the selection window caused by SP or the PP conferred by SMC.

Clinical studies investigating SMC’s impact on quintuple mutant spread have shown contradictory results.^13^ Mathematical models have assessed the effect of IPT on resistance spread in perennial settings.^14–19^ However, to our knowledge, no model has assessed SMC’s impact on the spread of quintuple mutants, nor investigated factors driving their spread. Moreover, it remains uncertain how the spread of quintuple mutants will reduce SMC’s effectiveness against clinical malaria. Here, we used an individual-based model of malaria transmission, OpenMalaria,^20^ combined with explicit parasite dynamics and drug pharmacokinetic and pharmacodynamic models, to address these questions. We assessed the rate of spread of the quintuple mutant and systematically quantified which factors drive its spread under various deployment strategies and seasonality settings. Finally, for a range of assumptions, we estimated the potential decrease in the effectiveness of SMC against clinical malaria attributable to the spread of the quintuple mutant.

## Research in context

### Evidence before this study

In March 2021 we performed a literature search for modelling studies that assessed the impact of SMC deployment on the spread of SP-resistant malaria parasites. PubMed was used with the keywords: malaria AND model* AND resistance AND seasonal malaria chemoprevention OR intermittent preventive treatment (IPT). We found six studies focused on the impact of IPT on the spread of resistance but no studies focusing on the impact of SMC on resistance spread. These studies assumed that the drug was continuously administered to the targeted population. Thus, it remains unknown how SMC implementation strategies (such as the number of cycles administered per year and the target age group(s)) affect quintuple mutant spread in seasonal settings. Four of these six studies assumed that parasites were fully IPT-drug resistant, and did not model partial resistance, as is the case for the quintuple mutant. Consequently, these studies ignored SP’s important residual prophylactic effect on the quintuple mutant. Five of the six studies assumed that the resistant genotype was resistant to both drug regimens used for IPT and first-line treatment. This assumption does not correspond to the use of SP for SMC in the Sahel where different first-line treatments are used (mainly artemether-lumefantrine).

### Added value of this study

To our knowledge, our study is the first to estimate the potential rate of spread of SP-resistant quintuple mutants under selection by SMC with SP+AQ. Our mechanistic model captures SP’s residual prophylactic effect on quintuple mutants, as informed by previous studies, and allows us to estimate its spread under SMC deployment for a range of settings. Clinical and observational studies remain the gold-standard evidence, however, evidence on the rate of spread of SP resistance and why it spreads is scarce. The added value of our study, informed by SP pharmacokinetics (PK) data and limited clinical evidence on quintuple pharmacodynamics (PD), is that we can extrapolate and explore dynamics of SP resistance spread under pressure from SMC to a large range of different epidemiological and clinical settings. For the first time, we estimate the dynamics of SP resistance spread and determine that a relatively long period is required to reach fixation (100% frequency) of the quintuple mutant. Moreover, our approach allows us to assess the protective effectiveness of SMC against malaria clinical cases in a parasite population composed solely of quintuple mutants.

### Implications of all available evidence

We found that the rate of spread of the quintuple mutant with partial resistance to SP is relatively slow if the target population for SMC is under ten or five years of age, and this rate strongly depends on the implementation strategy of SMC with SP+AQ. Our results support the continued implementation of SMC, as it will continue to prevent millions of clinical cases in Sahelian children despite the spread of the quintuple mutant. Our findings also highlight that in seasonal settings where the quintuple mutant is already highly prevalent, and clinical and severe malaria remains high, implementing SMC with SP+AQ may still have a marked clinical benefit.

## Methods

### Model calibration

OpenMalaria simulates *P. falciparum* dynamics in mosquitoes and humans,^21–23^ tracks multiple parasite genotypes, models their intra-host dynamics, and allows genotypes to have different sensitivities to drugs as specified in the pharmacokinetics and pharmacodynamics (PK/PD) model components.^24^ The model has been previously described in ^21–24^ and is briefly described in appendix p 4.

Using this model, we tracked quintuple mutant spread in a parasite population composed of quadruple mutants attributable to implementation of SMC with SP+AQ. For simplicity, we assumed that the quadruple mutant is the only competitor of the quintuple mutant because it is the most important competitor of the quintuple mutant for two reasons. First, the quadruple mutant is highly prevalent in the Sahel.^4, 9, 10^ Second, there is no apparent fitness cost associated with resistance to SP ^9, 25^ implying that all other genotypes (sensitive, single, double, and triple mutants) behave similarly to the quadruple mutant in individuals who did not receive SMC, although the quadruple mutant can develop a successful blood-stage infection a few days earlier than these genotypes in children who received SMC. As the quintuple mutant is already present in the Sahel, we focus on the spread of this genotype to high frequency and ignore denovo mutation, which will have a negligible impact on the spread of the quintuple mutant to high frequency.

We deployed SMC in two archetypal seasonality settings for a range of parasite prevalence in two to ten year-olds (*Pf*PR_2-10_) modelled via inputs with a range of entomological inoculation rates (EIR), in which approximately 85% of transmission occurs over three months (reflecting high seasonality, such as in Senegal) or four months (moderate seasonality, such as in Burkina Faso) (appendix p 5 figure S1.1). SMC in high seasonality settings was deployed three times monthly during the transmission season (as typically implemented in practice), or four times, with the additional cycle administered before or after the typical deployment period (appendix p 5 figure S1.2). Similarly, SMC in moderate seasonality settings was deployed four times monthly during the transmission season or five times, with the additional cycle administered before or after the typical deployment period (appendix p 5 figure S1.3). We deployed SMC to two different target age groups, children three months to five years (as typically implemented in practice), or three months to ten years. For each strategy, we simulated scenarios in which SMC coverage was constant or was decreased by 10% from the previous cycle (e.g., if cycle one is 80%, then subsequent cycles are 72%, 65%, etc.).^4, 11^

At each SMC cycle, children received one dose of SP and three daily doses of AQ, dosed according to their age (appendix p 6 table S1.1).^2, 26^ and were assumed to fully adhere to the regimen. We used two two-compartment PK/PD models with first-order absorption (appendix p 12-16 tables S1.4 and S1.5) to model AQ and its active metabolite. We assumed that AQ and its metabolites were effective against both mutants and provided a 17-day PP. SP is a synergistic two-drug combination making it challenging to simulate across a population.^27^ There is currently no PD data that report the blood-stage activity of SP against either the quadruple or quintuple mutant. We, therefore, made some simplifications and represented SP as a single, long-acting drug, denoted SP*, with one-compartment PK and first-order absorption (appendix p 8-11 table S1.3). We estimated the relationship between the PP and the half-maximal effective concentration (EC50) of SP* in OpenMalaria (appendix p 8-11 figure S1.3) and calibrated the EC50 of each genotype to match the PP reported by the literature.^5–7, 11^ This parameterisation demonstrated the same key epidemiological properties as SP in our simulations, i.e. (a) the ability, in combination with amodiaquine, to clear existing infections irrespective of whether they are quadruple or quintuple haplotypes, (b) chemoprophylaxis for quadruple and quintuple mutations of 35- and 21-days post-treatment, respectively, as suggested from the literature (figure 1A).^5–7, 11^ Note that the 35- and 21-day PP were average values over the targeted population. However, we captured variations of PP among individuals. Our model tracks individually assigned weights for each age band (appendix p 7 table S1.2), and children with different weights receive the same dose according to SP+AQ dosage recommendations (e.g. children between 1 to 5 years old, appendix p 6 table S1.1). Thus SP* reached a lower maximum concentration (mg/kg) in older, heavier children and reached its minimum inhibitory concentration faster in older children, leading to a shorter prophylactic period than in younger children. Our calibration predicted that the quintuple, but not the quadruple mutant, could develop a blood-stage infection before the next SMC cycle (figure 1B), and we successfully replicated a randomised clinical trial (see appendix p 17-20).

Note that children who received SMC could still be infected and access first-line treatment. This treatment was an artemisinin-based combination therapy (ACT) effective against both mutants (see legend of table 1, and appendix p 21).

**Table 1:** Parameters and their ranges investigated in the global sensitivity analyses of the rate of spread of the quintuple mutant. The range of SMC coverage at the first cycle reflects that reported by the largest SMC implementation study in the Sahel.^4^ The entomological inoculation rate (EIR) range reflects setting varying from low to high malaria transmission intensity. SMC is not recommended in settings with malaria prevalence below 10% in children age 2 to 10 years. An EIR of below five inoculations per person and per year would result in a lower prevalence than 10% in settings with high access to treatment and therefore was not investigated. The range for access to first-line treatment includes low to high access to treatment to cover a large spectrum of health system strengths. Children in the targeted groups could still contract malaria and obtain first-line treatment (an ACT) through the formal health sector, the same as individuals not targeted by SMC. The modelled ACT combined dihydroartemisinin with a partner drug whose elimination half-life was varied in the sensitivity analysis and captured the range of half-lives of lumefantrine, piperaquine, and mefloquine (appendix p 21). The partner drug was assumed to not be SP or AQ following WHO recommendations.^2^ ACT was fully effective against quintuple and quadruple infections. A Latin Hypercube Sampling algorithm was used to sample from the ranges (appendix p 24). ACT=artemisinin-based combination therapy. AQ=amodiaquine. SMC=seasonal malaria chemoprevention. SP=sulfadoxine-pyrimethamine.

| **Determinant** | **Definition** | **Parameter range** |
| --- | --- | --- |
| **Coverage at the first cycle of SMC** | Percentage of individuals from the target age group that received SP+AQ during the first cycle of SMC (%) | [70,100] |
| **Entomological inoculation rate (EIR)** | Mean number of infective bites received by an individual during a year (inoculations per person per year) | [5, 500] |
| **Level of access to first-line treatment** | Percentage of symptomatic cases who received treatment within 14-days of symptom onset (%) | [10, 80] |
| **Half-life of the partner drug for first-line ACT** | Time for the drug concentration of the partner drug for first-line ACT to fall by 50% (days) | [6, 22] |

### Identification of factors increasing SP resistance spread

For each seasonality setting and SMC deployment strategy, we systematically varied and quantified the influence of epidemiological, PK/PD, health system, and deployment factors (table 1) on the quintuple mutant spread using global sensitivity analyses (appendix p 24-25).^24^ We estimated the spread of the quintuple genotype in our model through the selection coefficient, which measures the rate at which the logit of the resistant genotype frequency increases each parasite generation (appendix p 24).^24^ We used Sobol’s method of variance decomposition (appendix p 25), which allowed estimation of first-order indices for each factor, which represent their influence on the spread. The 25^th^, 50^th^, and 75^th^ quantiles of the predicted rate of spread were reported for each parameter range. To illustrate results in a given time frame, we translated the selection coefficient to the time needed for the quintuple mutant to spread from 1% to 50% of inoculations, *T_50_*, for a set of parameter combinations that represent the Sahel (appendix p 34). We choose an initial frequency of 1%, since the frequency reported for the Sahel is around this value.^4^ Nevertheless, regions with an initial frequency above 1% would have a shorter *T_50_* (appendix p 34).

### Impact of the quintuple mutant on SMC effectiveness

To estimate the impact of SP resistance in the Sahel on SMC effectiveness, we estimated the protective effectiveness (PE) of SMC against parasite populations composed of 100% of the same genotype for which we varied the PP conferred by SMC with SP+AQ across simulations: 5-, 10-, 15-, 21- (mean PP against quintuple mutants), 25-, 30-, 35- (mean PP against quadruple mutants), and 42- (mean PP against sensitive parasites) days (appendix p 41 table S2.1). Note that when SP+AQ provided a PP lower than 17 days, we modelled some degree of resistance to AQ. We assessed the PE as the relative reduction in the incidence of clinical malaria in children three months to five years of age during the months of SMC implementation compared to the same population prior to SMC (appendix p 39-40).^4^ The PE was assessed for a set of parameter combinations (appendix p 39-40).

## Results

Global sensitivity analyses indicate that the levels of malaria transmission and SMC coverage consistently play a crucial role in the spread of the quintuple mutant over all SMC deployment strategies and seasonality patterns (figure 2A, appendix p 30 and 31 figures S2.5 and S2.6). Here we provide results for the deployment of four cycles of SMC per year at constant coverage to children under five years in the moderate seasonality setting (figure 2). High transmission levels increased the rate of spread. For example, when the EIR increased from 10 to 50 inoculations per person per year, the median selection coefficient increased by 64·0 (figure 2B). High SMC coverage levels also increased the rate of spread. For example, when the coverage increased from 70% to 80%, the median selection coefficient increased by 17·9% (figure 2C). The impact of coverage was stronger for SMC targeting children under 10 years versus those under 5 (figure 2A).

**Figure 2:**
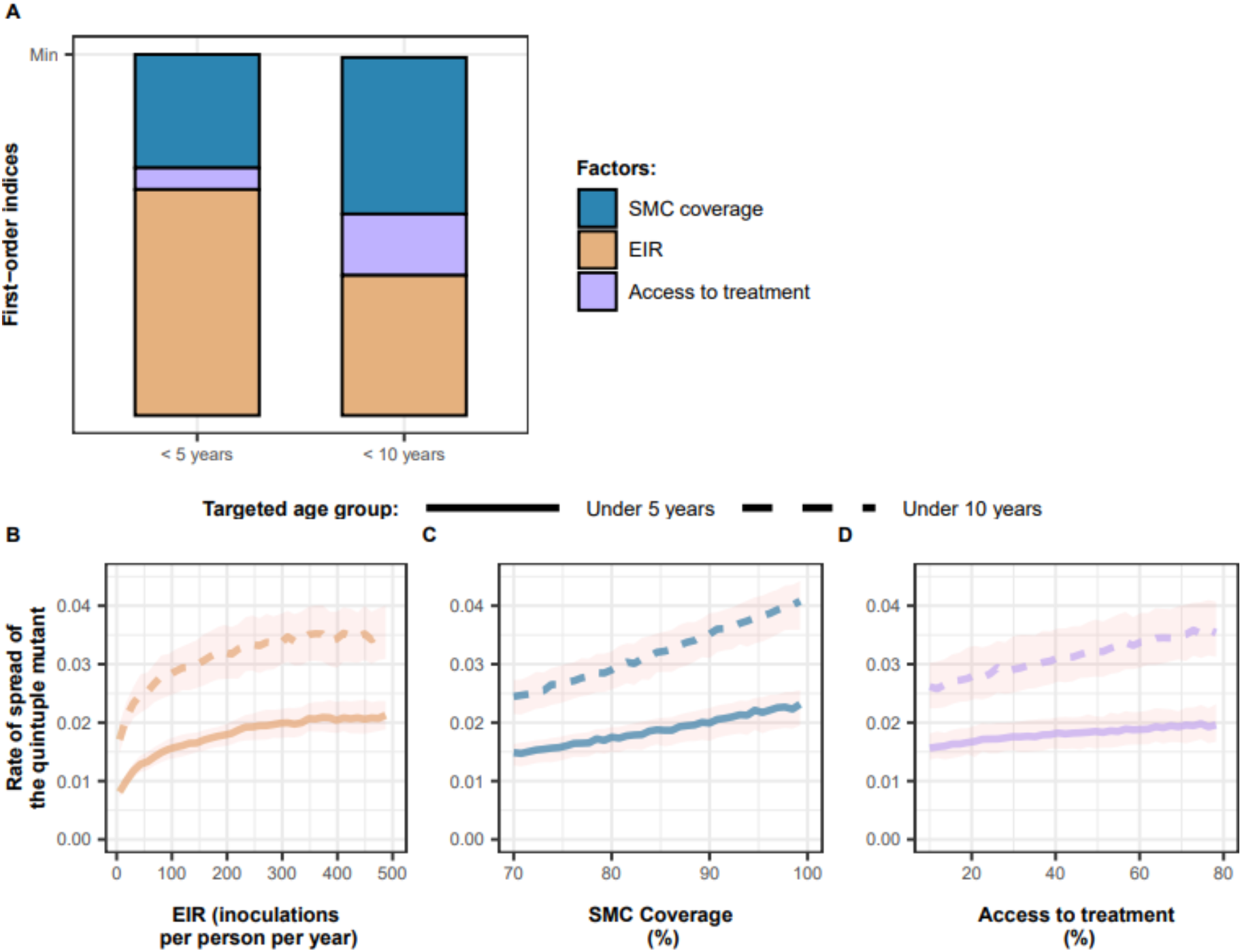
Influence of different factors on the rate of spread of SP-resistant quintuple mutants. (**A**) First-order indices of coverage (blue bar areas), EIR (orange), and level of access to treatment (purple) estimated during the global sensitivity analyses of the deployment of four cycles of SMC per year at constant coverage to children three months to five years of age or three months to 10 years of age in the setting with moderate seasonality. First-order indices for each factor are proportional to their area within the bars and represent their influence on the rate of spread. Parameter ranges explored were as follows: SMC coverage [70%, 100%]; EIR [5, 500] inoculations per person per year; and level of access to treatment [10%, 80%]. The PPs for the quadruple and quintuple mutants were 35- and 21- days, respectively. (**B**, **C**, **D**) Curves and shaded areas represent the median and interquartile range of the rate of spread (selection coefficient) of the quintuple mutant estimated from the global sensitivity analyses over the ranges of key parameters, (**B**) transmission level represented by EIR, (**C**) SMC coverage at cycle one, and (**D**) level of access to first-line treatment for symptomatic infections. The median and interquartile range were calculated during the global sensitivity analyses for deployment of four SMC cycles per year at constant coverage to children three months to five years (solid curves) or three months to 10 years (dashed curves) in a setting with moderate seasonality. The elimination half-life of the first-line ACT partner drug ([6, 22] days) is not displayed as it did not impact the rate of spread. Results for different seasonality settings, number of cycles deployed per year, and assumptions around SMC coverage reduction between cycles can be found in appendix p 30 and 31 figures S2.5 and S2.6. EIR=entomological inoculation rate. SMC=seasonal malaria chemoprevention.

The level of access to first-line treatment also slightly increased the rate of spread. Individuals not targeted by SMC can be infected by the quadruple and quintuple mutants, whereas individuals protected by SMC are more likely to be infected by quintuple mutants (appendix p 33 figure S2.8). Settings with higher access to treatment have significantly lower levels of parasite prevalence in individuals not targeted by SMC (above 5 or above 10 years old) compared to settings with lower access to treatment. However, children targeted by SMC are already largely protected by prophylaxis so the reduction in prevalence caused by improved access to treatment is small to negligible in this group (appendix p 32 figure S2.7 with SMC coverage of 95%). Thus, a rise in access to treatment disproportionally targets the quadruples in the non-SMC group, causing a slight increase in the frequency of quintuple mutants across the whole population (appendix p 33 figure S2.8), favouring its spread. For example, in settings with access to treatment of 30%, the estimated median selection coefficient increased by 12·4% (figure 2D) compared to a setting with access to treatment of 10%.

The predicted *T_50_* depends strongly on the SMC deployment strategy (figure 3, appendix p 35 figure S2.9). To illustrate this, we compared *T_50_* for various strategies in settings with medium transmission (EIR=75 inoculations per person per year, parasite prevalence of 40.3% in children 2-10 years prior to SMC, see appendix p 36 figure S2.10 for results related to parasite prevalence) and low access to treatment (25%), and an initial SMC coverage of 95% that decreased by 10% from each previous cycle (figure 3, red curves). With standard deployment of SMC (four cycles for children three months to five years) in settings with moderate seasonality, it took the quintuple genotype 53·1 years (95% CI 50·5– 56·0) to reach 50% of inoculations (*T_50_*). In contrast, it took 67·1 years (95% CI 63·2–71·5) with the standard SMC regimen (three cycles for children three months to five years) in highly seasonal settings. *T_50_* likely increases in high seasonality settings because it has one less SMC cycle, which reduces selection pressure.

**Figure 3:**
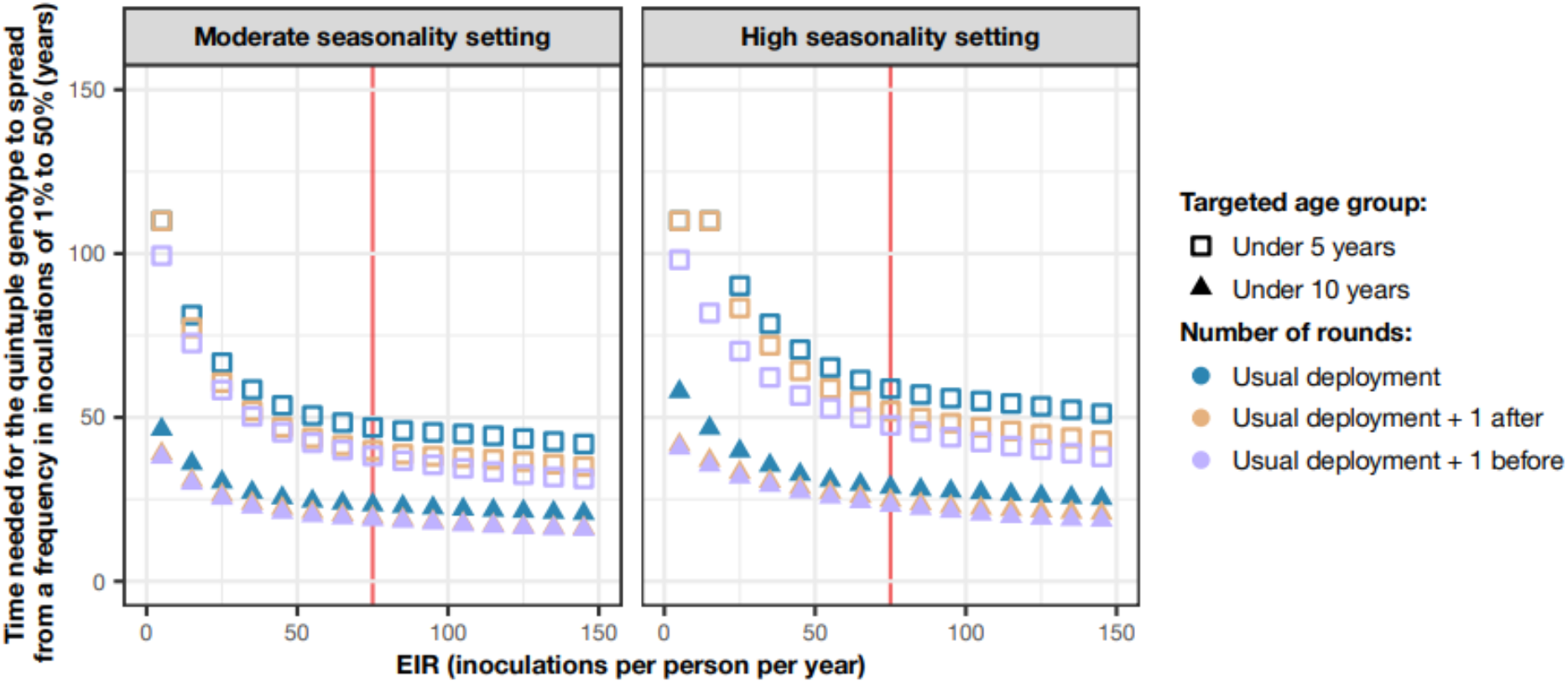
Predicted impact of SMC deployment strategies on spread of the SP-resistant quintuple mutant. Estimated time needed for the quintuple genotype to spread from 1% to 50% of inoculations, *T_50_,* when SMC was deployed to different age groups (children three months to five years (squares) or children three months to ten years (triangles)) in moderate and high seasonality settings with various numbers of cycles deployed per year (high seasonality: three (blue shapes) or four cycles with the additional cycle deployed before (purple) or after (orange) the current deployment period; moderate seasonality setting: four (blue) or five cycles with the additional cycle deployed before (purple) or after (orange) the current SMC deployment). *T_50_* was predicted for various transmission levels (ranging from 5 to 150 inoculations per person per year), assuming that SMC was deployed at an initial coverage of 95%, which was decreased by 10% from each previous cycle in a setting with high access to treatment (50%). Predictions at different levels of initial coverage and access to treatment can be found in appendix p 35 figure S2.9. Red lines highlight points of interest which are discussed in the results section.

In moderate seasonality settings, deploying an additional cycle of SMC at the beginning or end of transmission season decreased *T_50_* by approximately 13 and 10 years, respectively. Similarly, in high seasonality settings, deploying an additional cycle before or after the typical deployment reduced this time by 16 and 15 years, respectively.

Increasing the SMC target population from children aged three months to five years to children aged three months to ten years means almost twice the number of individuals received SMC, accordingly, the *T_50_* was halved. For example, in moderate transmission settings, *T_50_* decreased from 53·1 (95% CI 50·5– 56·0) to 26·4 years (95% CI 25·6–27·3) when administering four cycles of SMC to children under ten, compared with administering to children under five. Similarly, in high transmission settings, *T_50_* decreased from 67·1 (95% CI 63·2–71·5) to 35·9 years (95% CI 33·6–26·7).

As expected, the PE of SMC was strongly dependent on the length of the PP (figure 4, appendix p 42 figure S2.13). PE decreased with shorter PP, but still remained important in a parasite population composed only of quintuple mutants (appendix p 42 figure S2.13). For example, with standard SMC deployment with 95% coverage (decreasing by 10% from each cycle) in moderate seasonality, low access to treatment (25%) setting, SMC prevented 79·0% (95% CI 77·8–80·8) and 60·4% (95% CI 58·6–62·3) of malaria episodes across all transmission intensities when the parasite population was composed by 100% of quadruple mutants (resulting in a 35-day PP, figure 4, yellow box and shaded area) or 100% of quintuple mutants (resulting in a 21-day PP, figure 4, green box and shaded area), respectively. This suggests that SMC will retain some effectiveness even if the quintuple mutant becomes fixed (i.e. reaches 100% frequency) in the population.

**Figure 4:**
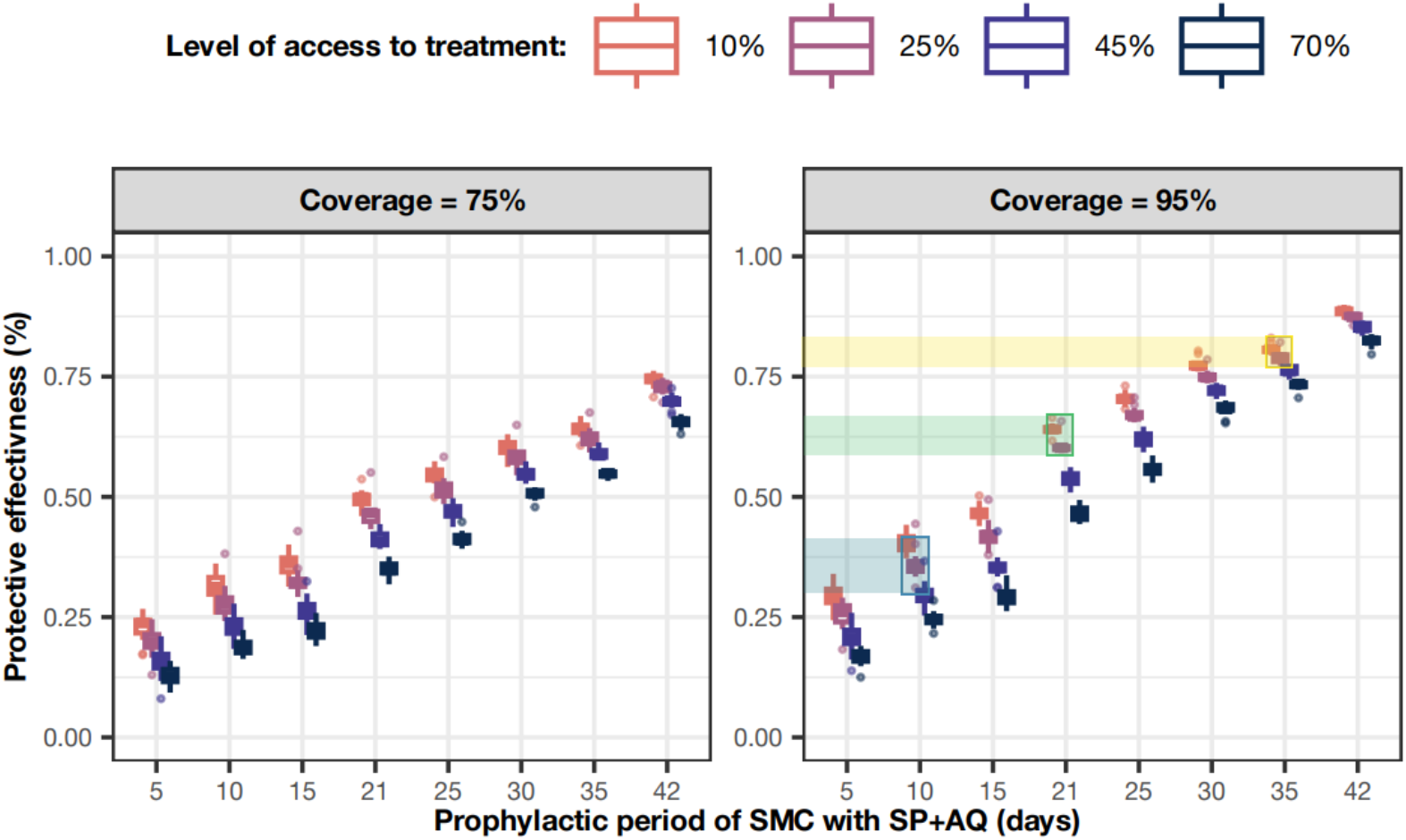
Protective effectiveness of SMC for a range of prophylactic periods. The protective effectiveness of SMC (the relative reduction in the number of clinical malaria cases in children aged three months to five years during the months of SMC implementation) when four cycles of SMC were delivered to these children at different coverage levels (75% and 95% with coverage decreased by 10% from each cycle), in settings with diverse transmission intensities (from 5 to 150 inoculations per person per year), and at levels of access to treatment from 10% (orange boxplot) to 70% (dark blue boxplot). The protective effectiveness was assessed against parasite populations composed of 100% of the same genotype for which we varied the PP conferred by SMC with SP+AQ across simulations: 5-, 10-, 15-, 21- (mean PP against quintuple mutants), 25-, 30-, 35- (mean PP against quadruple mutants), and 42- (mean PP against sensitive parasites) days (appendix p 41 table S2.1). Note that when SP+AQ provided a PP lower than 17 days, we modelled some degree of resistance to AQ. The x-axis is not equally spaced in order to illustrate the assumed PP against each genotype. The circles represent outliers. The results for different SMC coverage levels can be found in appendix p 42 figure S2.13. Boxes and shaded areas identify examples described above in the main text.

If a genotype acquires higher degree of resistance to SP at the same time as some degree of resistance to AQ, the prophylactic period conferred by SMC with SP+AQ will likely be shorter than 21 days. Current evidence suggests the PP of AQ against partially AQ-resistant genotypes to be slightly longer than 10 days.^12^ However, even if SP+AQ SMC provides a PP of 10 days, SMC could still prevent 36·0% (95% CI 34·5–37·5) of clinical cases for the same deployment and setting as above (figure 4, blue box and shaded area). In addition, for the same deployment, this resistant genotype would still require 26·5 years (95% CI 20·8–32·2) on average to spread from 1% to 50% frequency across settings with an EIR of 50 inoculations per person per year (appendix p 43 figure S2.14, blue box and shaded area).

## Discussion

To our knowledge this is the first modelling study to (1) estimate the impact of SMC implementation on the spread of the *dhfr/dhps* quintuple mutant resistant to SP, and (2) assess the impact of this spread on SMC’s ability to prevent *P. falciparum* malaria. In our model, the current implementation of SMC (four monthly cycles to children three months to five years assuming 95% initial coverage declining by 10% from each cycle) resulted in the relatively slow spread of the quintuple mutant in settings typical of the Sahel region, whereby the quintuple mutant takes over 50 years to spread from a frequency of 1% to 50% in settings with low access to first-line treatment. We further predicted that spread of the quintuple mutant could accelerate due to changes in SMC deployment, such as its deployment at higher coverage, adding additional cycles of SMC per year, and expanding the targeted age group. Nevertheless, the time to fixation was still relatively long. Our study further shows that SMC with SP+AQ will remain a valuable tool to prevent uncomplicated malaria throughout the next decade, even if its prophylactic period is likely to decrease over time with the slow spread of the quintuple mutant with reduced sensitivity to SP. Our model predicts that SMC will remain effective at preventing malaria morbidity even with a high frequency of quintuple mutations, for the typical SMC delivery, for example, preventing 60·4% (95% CI 58·6–62·3) of clinical cases in settings typical of the Sahel. Implementation of SMC in seasonal settings where the quintuple mutant is already highly prevalent should be considered. Its implementation could considerably reduce malaria-related morbidity in children at risk.

Multiple factors explain this relatively slow spread of the quintuple mutant. First, SMC is only deployed to a minority of individuals in the population, so only this minority could potentially select for the quintuple genotype. We found that SMC coverage was a critical factor influencing spread, since, unsurprisingly, the more individuals receiving SP+AQ, the greater the selection for the quintuple mutant. However, even at 100% coverage, when targeting children three months to five years, less than 20% of the population received SMC at each cycle. Second, SP creates only a short selection opportunity for the quintuple mutant among treated children. The selection window was equal to 14-days (i.e., 21- to 35-days post-SMC). The selection opportunity was reduced to 9-days (i.e., 21- to 30-days post-SMC) when children received a subsequent SMC cycle scheduled at day 30 post-SMC. Finally, individuals who receive SMC and become newly infected by the quintuple mutant can have their infection cleared by the AQ component in the next cycle, further limiting spread. Our findings for slow spread of the quintuple mutant are in agreement with previous studies, which reported that SMC leads to a slow or no marked increase in the quintuple mutant’s frequency.^4, 13, 28^

We demonstrated that deploying SMC to children aged three months to ten years compared with that for children three months to five years almost doubled the rate of quintuple mutant spread. This is presumably because the number of individuals receiving SMC approximately doubles. The relative importance of SMC coverage on the rate of spread was more important when we expanded the target age group, as a percentage increase in coverage led to incrementally more children receiving SMC in total. In addition, adding one additional cycle of SMC per year reduced the time needed for the quintuple mutant to reach 50% of inoculations by approximately ten years. Nevertheless, previous studies have highlighted that extending SMC to children under ten years or adding extra cycles of SMC per transmission season could provide substantial health benefits.^11, 26^

We show that spread of SP resistance increases with higher transmission intensities, likely due to several reasons. First, in a high transmission setting with a higher force of infection, the quintuple mutant may be able to circulate faster in the population. Second, there are higher levels of acquired immunity in high transmission settings. Suppose a quintuple mutant becomes established in the selection window in high transmission areas. In that case, it is more likely to be asymptomatic and hence remain untreated by first-line treatment, and thus it may spread faster. However, in real life, it is more complex to assess the frequency of quintuple mutants in high-prevalence settings due to the higher number of patients infected with multiple parasites. Thus, it is essential to ensure efficient surveillance of molecular markers in these areas.

We observed that SMC retains a substantial level of PE even if the quintuple mutant spreads to 100%. This is because SP inhibits development of successful blood-stage infection of the quintuple mutant for 21-days post-treatment, and children receive SP+AQ every 30-days, so children are protected during most of the SMC deployment period. In addition, children that develop blood-stage infections after 21-days have their infections cleared by AQ in the next cycle of SMC. Consequently, SMC will remain a valuable tool to reduce malaria morbidity in the Sahel despite the spread of the quintuple mutant. Critically, these findings suggest that SMC with SP+AQ could be implemented in seasonal regions where the quintuple mutant is already prevalent, such as in southern and eastern Africa. Currently, SMC with SP+AQ is not implemented in these regions due to the high prevalence of the quintuple mutant.^2^ We argue that further evidence is needed to challenge or confirm our predictions. Results of a recently published SMC trial with SP+AQ in Uganda, where the quintuple mutant is more prevalent, confirm our findings.^29^

Our recommendations and modelling results depend on several assumptions. First, given limited data on SP’s mode of action and synergism, we used existing clinical data to inform a model of SP as a long-acting drug providing a PP of 21-days on average against the quintuple mutant and 35-days on average against the quadruple mutant.^5, 6, 11^ This approach allowed us to capture the PP provided by SP and the selection window. Our approach captured the average population level PP of SP against the quintuple mutant of 21 days and the variation of PP among individuals due to differences in weight and dosage among children. However, we did not model individual variability in PK parameters. PK variability would cause faster drug concentration decay and shorter PP in some individuals than others but will not strongly impact the average rate of spread in the population. In addition, our reference parameterisation of SP depends on limited data on the PP of SP in regions where the quintuple and quadruple mutants are prevalent.^5, 6, 11^ Our additional analysis on this PP, shows that if the PP against the quintuple mutant is shorter than 21-days, the spread of resistance will be faster (appendix p 43 figure S2.14). It is important to note further that AQ provides a blood-stage PP of 17-days against AQ-sensitive parasites.^12^ Thus, SMC would still offer a PP for 17-days primarily driven by AQ. Consequently, our estimation of the rate of spread and the effectiveness of SMC would not change dramatically unless parasites also acquire resistance to AQ, for which the PP conferred by AQ would still be longer than 10-days.^12^ This also means that if a parasite more resistant to SP emerges, such as the sextuple mutant (with an additional mutation: *dhps*-A581G) as observed in a few settings in East Africa,^9^ we would still expect limited spread and the protective effectiveness of SMC to be retained thanks to AQ.

Second, we assumed that both genotypes were sensitive to AQ and that children were fully adherent to the three doses of AQ.^2^ If we had assumed that resistance to AQ is present in the population or adherence to AQ is low, the PP conferred by SMC against each mutant should not change as SP determines the length of this period. However, a high degree of resistance to AQ or low adherence to AQ could lead to treatment failure of SP+AQ, which would increase the spread of the quintuple mutant as the rate of treatment failure would probably be higher for the quintuple than the quadruple mutant. Nevertheless, only markers for low degrees of resistance to AQ have been observed in the Sahel (*Pfcrt-CIVET* + *pfmdr1-86 Tyr* + *184 Tyr*) at low and declining prevalence (0·5% of samples from 2018).^4^ Thus, these mutations should not impact the spread of the quintuple mutant. Considering the potential interaction between AQ and artemisinin resistance, further modelling studies should assess how SMC implementation may impact the spread of partial artemisinin resistance.

Fourth, for simplicity, we assumed that the quadruple mutant is the only competitor of the quintuple mutant, as it is currently the most important competitor. We effectively modelled that sensitive parasites, single, double, and triple mutants behave similarly to the quadruple mutant in individuals who did not receive SMC, as we assumed no apparent fitness cost associated with SP-resistance, as suggested in a previous study.^9, 25^ Nevertheless, our estimates of time to fixation of the quintuple mutant will be on the lower end, as we have disregarded some competition factors that would increase this period.

Fifth, we did not model the potential effect of pyrimethamine on the liver-stage of *P. falciparum*. Previous studies suggest that the liver-stage effect of pyrimethamine is reduced against quadruple and quintuple mutants, as they have three mutations conferring resistance to pyrimethamine.^30^ Thus, we may have underestimated (rather than overestimated) the effectiveness of SMC, as we did not model these liver-stage effects. However, this assumption does not affect our estimation for quintuple mutant spread, as the liver-stage effect of pyrimethamine is similar for both genotypes.

Lastly, we focused on the spread of the quintuple mutant favoured by SMC. However, in the real world, SP’s use in the private sector or other interventions, such as intermittent preventive treatment in pregnancy,^3^ could also favour the spread of the quintuple mutant. In addition, in the near future, other interventions can be deployed, such as vector-based interventions, that could further impact the rate of spread. This should be further assessed.

In conclusion, our assessment of the risk of spread of the quintuple mutant and associated consequences are reassuring overall, but should be validated by other modelling studies and, importantly, through clinical trials or implementation studies. However, mutants with a high degree of resistance to SP and AQ could emerge at any time in the Sahel. Therefore, routine molecular surveillance alongside efficacy testing for detected mutants must continue.

## Data Availability

Individual participant-level data was not used in this study. Parameter values used to inform the model were extracted from the literature as described in the main text or in the appendix. All data and code used to produce the figures are available at https://zenodo.org/record/7244708. In addition, the code used to run the simulations and perform the analyses can be found at https://zenodo.org/record/7244732. The individual-based model of malaria transmission and epidemiology used in the study has an open-access code (https://github.com/SwissTPH/openmalaria) and documentation (https://github.com/SwissTPH/openmalaria/wiki).

## Contributors

MAP conceived the study. TM, TL, IMH, and MAP designed the study. TM led and performed the literature searches to parameterise the model and performed the analyses. MAP further verified the model and analysis. TM, TL, IMH, SLK, and MAP interpreted results and examined their implications. TM wrote the first draft of the manuscript and created the tables and figures. TL, SK, IMH, and MAP revised the manuscript. All authors reviewed and approved the final manuscript. MAP acquired the funds and managed the project.

## Acknowledgments

This study was funded through a Swiss National Science Foundation Professorship grant of MAP (P00P3_170702). TL was funded through a Marie Curie Individual Fellowship (839121, Horizon 2020). Calculations were performed at sciCORE (http://scicore.unibas.ch/) scientific computing centre at the University of Basel. This article represents the views of the authors and not necessarily those of the funders.

## Supplementary Materials

### 1 Model description and parameterisation

#### 2 Individual-based model of malaria transmission dynamics

We parameterised and used our existing individual-based model of malaria transmission, OpenMalaria, that simulates the dynamics of *Plasmodium falciparum* in mosquitoes and humans^1–3^ as described previously.^2–5^ Here, we briefly described the main attributes of the model with a focus on features that allowed us to model drug resistance.

OpenMalaria is an ensemble of model components that is run with a time step of 5 days. The user can specify multiple parasite genotypes with different degrees of drug sensitivity and initial frequencies in the population. The human population size and age structure are constants throughout the simulations. Each individual human in the population was separately tracked from birth to death. When an individual becomes infected, within-host parasite dynamics are simulated with one of the model components.^6^ The model simulates parasite densities based on the parasite multiplication rate and the effect of innate, variant, and acquired immunities, which reduce the multiplication rate.^6^ An immunity profile for each individual is developed based on their cumulative parasite exposure.^7^ Infants are also protected by maternal immunity during the first months of life.^7^ The within-host model allows for concurrent infection of multiple parasite genotypes within the same host and captures that different parasite genotypes indirectly compete within-hosts as immunity regulates the total parasite load. Users may also specify a reduction of the within-host multiplication rates for a particular genotype to capture a potential fitness cost associated with mutations that differ between genotypes. The simulated parasite densities influence diagnostic outcomes, patient symptoms,^1^ risks of severe malaria and death,^8, 9^ and the probability that a feeding mosquito becomes infected.^10^ The probability that an infection is symptomatic depends on an individual’s levels of innate immunity (essentially a pyrogenic threshold that is also altered by exposure) and acquired immunity (their cumulative parasite exposure).^11^

A case management component describes treatments of uncomplicated and severe cases based on the previous use of drugs to treat the same episode, as well as levels of access to health services.^12^ In addition, the model allows to implement drug-based interventions in the population at specific timings, such as for seasonal malaria chemoprevention (SMC).

Pharmacokinetic (PK) models within OpenMalaria (with one-, two- or three-compartments) track drug concentrations in individuals who have received drugs.^13–15^ The effect of drug concentration on the within-host parasite multiplication rate is estimated using a pharmacodynamics (PD) model.^13–15^ Pharmacodynamics parameters can be parameterised individually for each genotype to model different degrees of drug susceptibility.^4^

Malaria parasite transmission in mosquitoes is modelled with a periodically forced deterministic model component to capture vector feeding behaviour and track the infectious states of the mosquitoes.^2^ The model periodicity allows to capture of seasonality transmission patterns. The different parasite genotypes are followed in the mosquitoes, but recombination is not considered (recombination would have no effect in this study as the two mutants considered differ by only one mutation). Numbers of newly infected human hosts depend on the simulated entomological inoculation rate (EIR). The genotype for new human infections is randomly drawn from genotype frequencies in humans infections from the previous five time-steps. Then liver-stage immunity reduces the number of infections reaching the blood-stage. Note that to track the rate of spread of the quintuple mutant, we estimate the frequency of the quintuple mutant across all inoculations (see section 2.1).

#### 3 Parameterisation of SMC

We modelled SMC as the monthly deployment of sulfadoxine-pyrimethamine (SP) plus amodiaquine (AQ) in target age groups during the transmission period. As explained in the main article, we deployed SMC to different age groups (between 3 months and five year-olds or between 3 months and ten year-olds) in diverse seasonality settings (figure S1.1) with various numbers of cycles deployed per year (figure S1.1). Children under five years received the dosage regimen of SP+AQ recommended by the World Health Organization (WHO) (table S1.1).^16^ Children under ten years received the AQ+SP dosage regimen used in Senegal (table S1.1).^17^ In the following sub-sections, we describe how we modelled SP and AQ using PK/PD models. In addition, we show how our parameterisation could replicate the results of a randomised non-inferiority trial comparing dihydroartemisinin-piperaquine with SP+AQ.^18^

**Figure S1.4:**
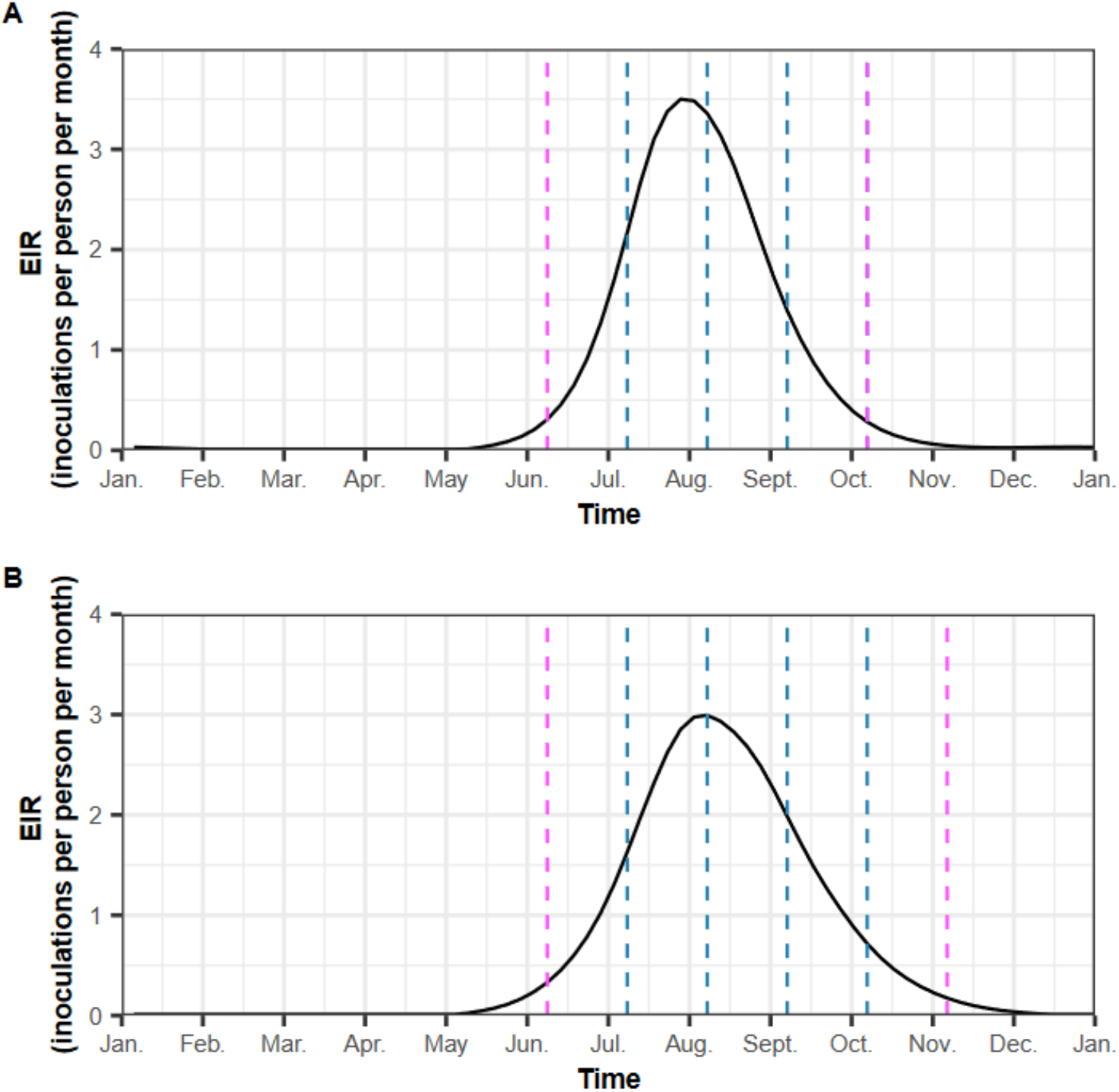
Seasonality pattern of transmission and timing of SMC deployment. The figure illustrates the entomological inoculation rate (EIR, in inoculations per person per year) across time in (A) high and (B) moderate seasonality settings. In high seasonality settings, SMC was deployed three times per year (as typically implemented) or four times per year, with the additional cycle administered before or after the typical deployment period. Similarly, for moderate seasonality settings, SMC was deployed four times per year (as typically implemented in this type of setting) or five times per year, with the additional cycle administered before or after the typical deployment period. Blue dashed lines represent standard SMC deployment. Pink dashed lines represent additional cycles of SMC that were either deployed before or after the standard deployment.

**Table S1.2:**
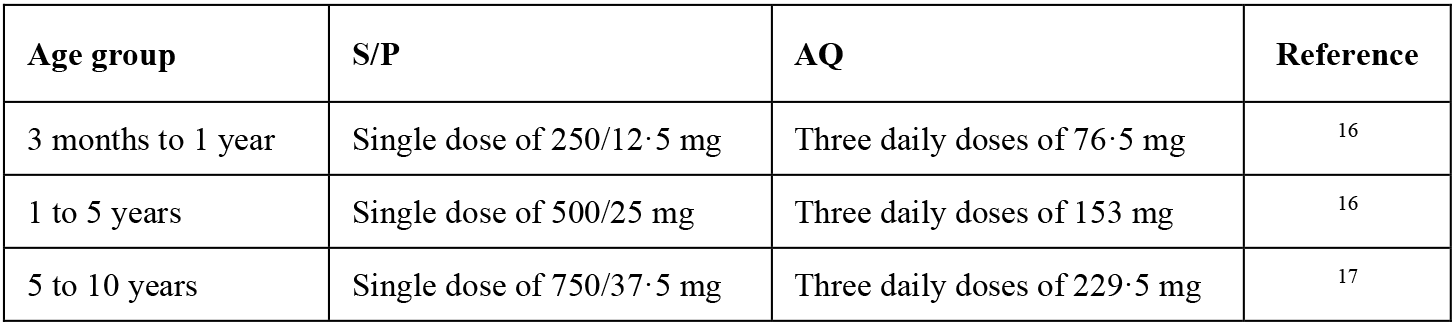
Dosage regimens of sulfadoxine/pyrimethamine and amodiaquine by age group. S/P=sulfadoxine/pyrimethamine. AQ=amodiaquine

**Table S1.3:**
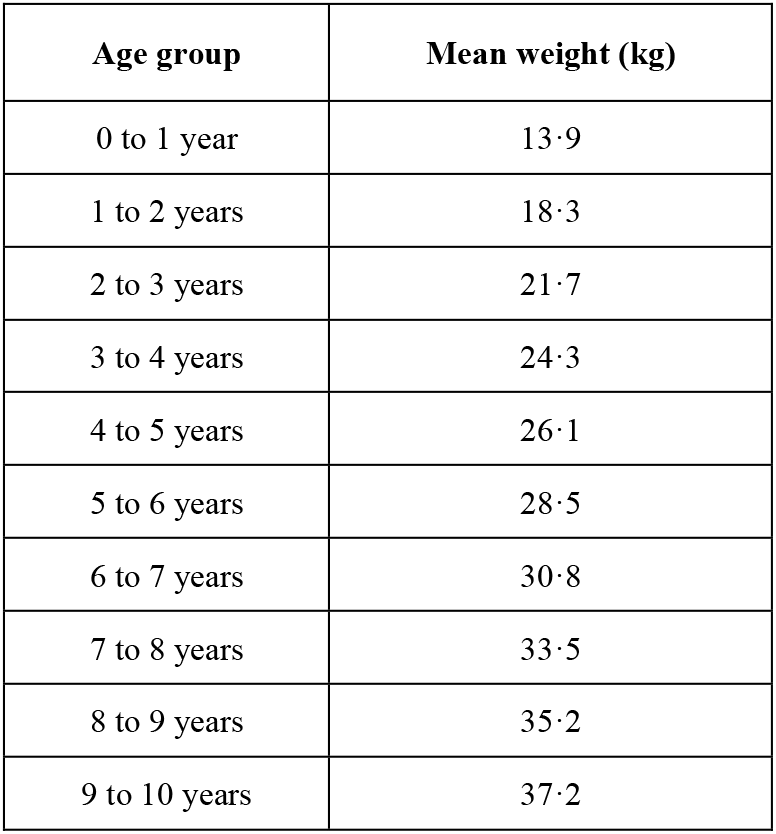
The mean weight of individuals by age bands.

##### 3.1 Sulfadoxine and pyrimethamine

Sulfadoxine and pyrimethamine have a synergic effect on the blood-stage of *P. falciparum*. This synergic effect is not well understood and is challenging to capture in PD models.^19, 20^ For simplicity, we modelled the two drugs as a single long-acting drug with a similar duration of action as the expected synergetic combination of sulfadoxine and pyrimethamine. This long-acting drug was denoted as ‘SP*’, and modelled using a one-compartmental model with first-order absorption and parameterised with values used by de Kock *et al.*^21^ to model sulfadoxine. We parameterised the drug with PK parameters for sulfadoxine (table S1.3) as it has a longer half-life than pyrimethamine. Thus, we captured the maximum time until one SP drug component was present at active concentrations in an individual. We compared our model prediction with the predictive check from De Kock *et al.*^21^ (figure S1.2). Our predictions centre around the median predictive check from De Kock *et al.*^21^. Note that we predicted less variation in the concentrations of sulfadoxine between individuals than De Kock *et al.*^21^ (see Figure 1 from de Kock *et al.*^21^). These differences are because our model does not include (1) individual variations for PK parameters, (2) variations in the drug clearance rate due to maturation, and (3) variations in drug bioavailability due to malnutrition, as was done in the study by de Kock and colleagues.^21^ However, the concentration of SP* still varies among individuals in our model since we specify a weight for each age band (table S1.2), and children with different weights receive the same dose according to SP+AQ dosage recommendations (e.g. children between 1 to 5 years old, table S1.1). Thus older, heavier, children receive a lower dose in terms of mg/kg and reach lower concentrations faster than younger, lighter children. This introduces variability in the prophylactic periods (PP) between children enrolled in SMC, as would be expected in clinical settings.

For the PD model, the maximum killing rate was parameterised based on an estimate of the synergetic maximum killing rate of SP (table S1.3).^19^ As informed by our literature review (described in the main article), we modelled that quadruple and quintuple mutants could develop a successful blood-stage infection after 21 days and 35 days on average, respectively.^22–25^ The length of the prophylactic period varied among individuals due to variations of SP* concertation. As described above, older children reach lower concentrations faster than younger children and thus experience shorter prophylactic periods. To achieve this, we parameterised that the quintuple mutant had a higher half-maximal effective concentration (EC50) than the quadruple mutant to capture that the quintuple mutant was less sensitive to low drug concentrations. To determine the EC50 for each mutant, we established the link between the average prophylactic period and the EC50 for SP*. This was done by simulating deployment of one cycle of SMC with only SP* to all children aged three months to five years, with full coverage (equal to 100%) in a non-seasonal setting in which these children had no access to first-line treatment. We ran multiple simulations over which we varied SP*’s EC50 and transmission levels (EIR=5, 50, 100, and 150 inoculations per person and per year). Each simulation started with a burn-in phase of 130 years, using ten seeds for 100,000 humans. For each simulation, we estimated the prophylactic period of SP* as the median time before detecting blood-stage infection in children during a follow-up period of 42 days after SMC deployment in alignment with Desia *et al*.^24^ Blood-stage infections could develop either due to treatment failure from a previous infection or reinfection reflecting real-life scenarios.^24^

As expected, the prophylactic period decreased with higher EC50 (figure S1.3). We found that the relationship between EC50 and the prophylactic period was stable in settings with EIR values ranging from 50 to 150 inoculations per person per year. However, for similar EC50 values, the prophylactic period was longer at an EIR of five inoculations per person per year than at higher transmission intensities. This result was because children needed more time to be reinfected in low transmission settings than in high transmission settings. Studies that assessed the prophylactic period of SP against the quintuple and quadruple mutants occurred in settings where EIR ranged from 50 to 150 inoculations per person per year. In these settings, SP* needed an EC50 of 2·39 mg/µl against the quadruple mutant and an EC50 of 24·2 mg/µl against the quintuple mutant to provide the wished prophylactic periods (figure S1.3).

In summary, we reduced the complexity of SP treatment (i.e., a 2-drug combination with synergistic killing) to a single “drug” based on SP pharmacokinetics. This simplification allowed us to recover the key pharmacological factor in our simulations, i.e., periods of prophylaxis following treatment for the two genotypes (quadruple and quintuple mutants) (figure 1A from the main text). This subsequently enabled us to predict the two key outcomes for our study, (1) the clinical impact of the quintuple mutant when it replaced the quadruple mutant (e.g., figure 1B from the main text), and (2) generates the window of selection which determines the rate at which the quintuple mutant replaces the quadruple haplotype (figure 1A from the main text). Note that neither genotype encodes resistance to treatment in our model, as the AQ component of SP+AQ used in SMC is still effective, nor does ACT used in formal health service delivery reliably clear parasite infection.

**Table S1.4:** PK/PD parameters for SP*. EC50=half-maximal effective concentration. SP*= the single long-acting drug with a similar duration of action as the expected synergetic combination of sulfadoxine and pyrimethamine.

| Parameter | Value | Reference |
| --- | --- | --- |
| Volume of distribution (l/kg) | 0.29 | <sup>21</sup> |
| Absorption rate (per day) | 12.50 | <sup>21</sup> |
| Elimination rate (per day) | 0.12 | <sup>21</sup> |
| Maximum killing rate (per day) | 2.30 | <sup>19</sup> |
| EC50 (mg/l) of the quadruple mutant | 2.40 | Estimated |
| EC50 (mg/l) of the quintuple mutant | 24.00 | Estimated |
| Slope of the effect curve | 2.10 | Assumed |

**Figure S1.5:**
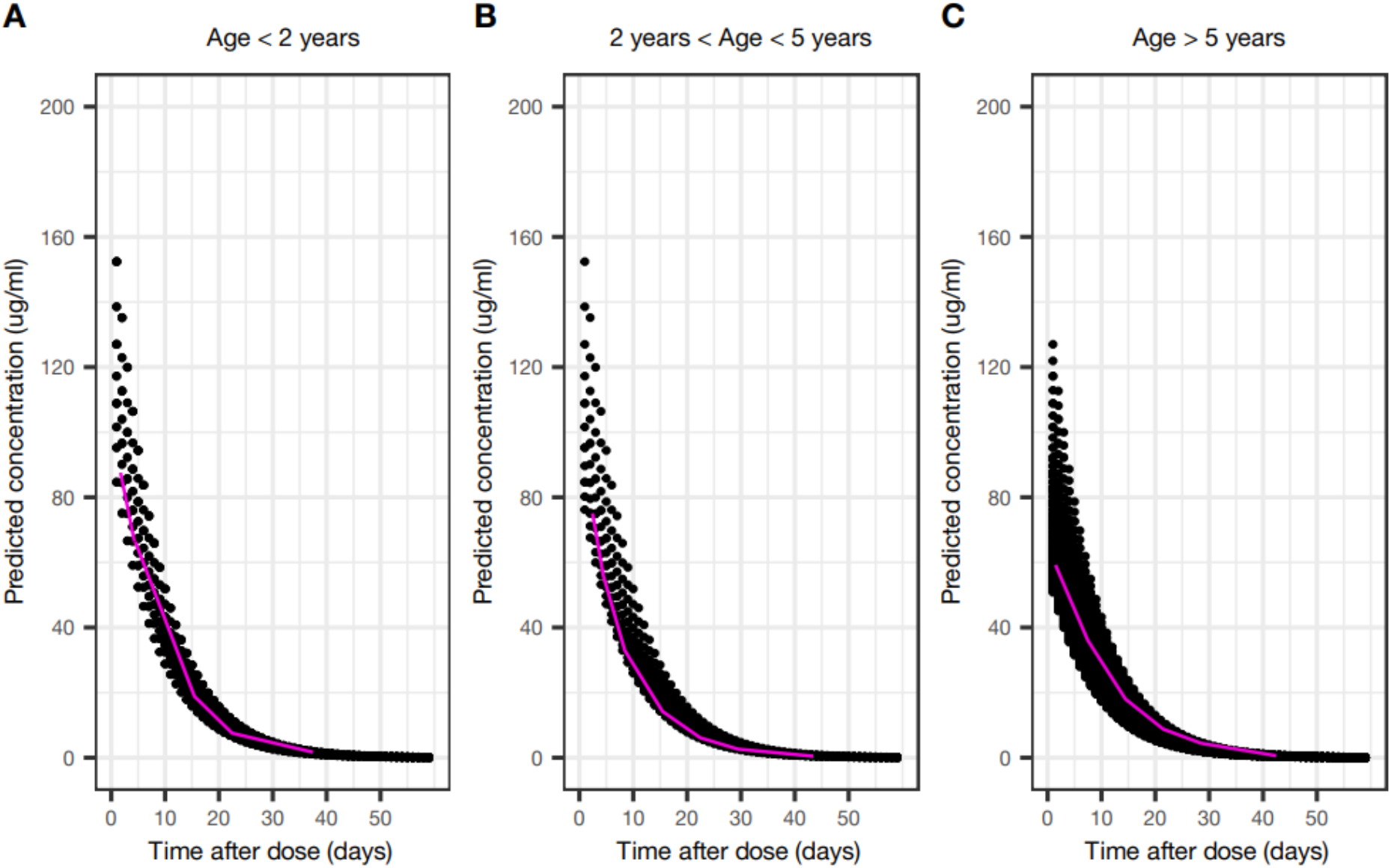
Comparison between sulfadoxine concentrations predicted by de Kock *et al.*^21^ and those from our model. Black dots represent the concentrations of sulfadoxine predicted by our model for individuals (A) under two years of age, (B) between two and five years of age, (C) and older than five years of age. Pink lines denote median concentrations from the predictive check from de Kock *et al.*^21^. Pink lines were drawn based on figure 1 from de Kock *et al.*^21^

**Figure S1.6:**
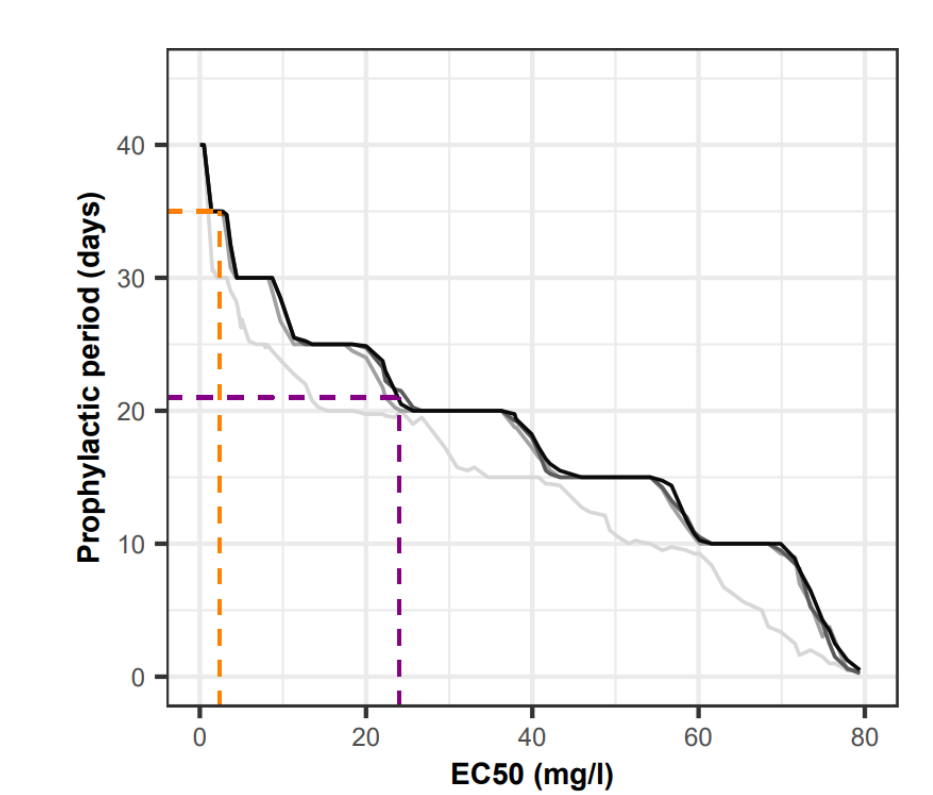
Predicted relationship between EC50 and the prophylactic period conferred by SP* across various transmission settings. The figure illustrates the predicted relationship between the EC50 and the prophylactic period conferred by SP* in perennial transmission settings with EIR equal to 5 (light grey curve), 50 (grey curve), 100 (dark grey curve), and 150 (black curve) inoculations per person per year. The purple and orange dashed lines represent the estimated EC50 for the quintuple and quadruple mutants, respectively. SP*= the single long-acting drug with a similar duration of action as the expected synergetic combination of sulfadoxine and pyrimethamine.

##### 3.2 Amodiaquine

In the human host, AQ is converted into its active metabolite, desethylamodiaquine (DEAQ). Our individual-based model does not include a conversion PK model with multiple compartments. Therefore, we modelled AQ and DEAQ as two separate drugs as in Hong *et al.*^26^. Each drug component was modelled with a two-compartment model with first-order absorption (Tables S1.4 and S1.5). We captured the fast metabolism of AQ into DEAQ by assuming that both drugs had a similar dosage, absorption rate, and that AQ had a short elimination half-life. We compared our model prediction with those from a more complex model applied by Ali and colleagues^27^ that was fitted to 261 individuals.^27^ Their model simulated AQ using a two-compartment model and captured AQ metabolism into DEAQ, which in turn is described by a three-compartment disposition model.^27^ *Ali et al. (2018)* used a dosing regimen based on the weight of individuals (individuals weighing between 4 to 8 kg, between 9 to 17 kg, between 18 to 35 kg and above 36 kg received 67.5 mg, 135 mg, 270 mg and 540 mg of AQ, respectively.^27^ We used the same dosage regimen as Ali *et al. (2018)* to replicate their figure and confirm the accuracy of our methodology, namely recover plasma concentrations given AQ dosages above. Note, apart from the validation, our study uses the dose regimen reported in Table S 1.1, matching standard dosing by age as recommended by WHO.^16^ We obtained similar DEAQ concentrations on day seven of the simulations (figure S1.4) and maximum DEAQ concentrations reported by Ali *et al.* (figure S1.5).^27^ Note that we tended to underestimate the maximum concentration in the heaviest weight groups compared with Ali *et al.*^27^. These differences arise because our model does not include variation in clearance rates based on weight and age of the children as done by *Ali et al.*^27^. In addition, we predicted less variation for day seven and maximum DEAQ concentrations across individuals than in the original figures 4A and 4C of Ali *et al.*^27^ (which also show interquartile ranges), because our model does not include individual variations of PK parameters as done by *Ali et al.*^27^. Finally, the PD parameters of AQ and DEAQ were taken from model parameters published by Hong *et al.*^26^

**Table S1.5:** Amodiaquine PK/PD parameters. EC50=half-maximal effective concentration.

| Parameter | Value | Reference |
| --- | --- | --- |
| Absorption rate (per day) | 2·16 | <sup>26</sup> |
| Elimination rate (per day) | 42·00 | <sup>26</sup> |
| Rate at which the drugs move from the central compartment to the peripheral compartment (per day) | 51·00 | <sup>26</sup> |
| Rate at which the drugs move from the peripheral compartment to the central compartment (per day) | 1·46 | <sup>26</sup> |
| Volume of distribution (l/kg) | 8·00 | <sup>26</sup> |
| Maximum killing rate (per day) | 2·30 | <sup>26</sup> |
| EC50 (mg/l) | 0·0043 | <sup>26</sup> |
| Slope of the effect curve | 7·00 | <sup>26</sup> |

**Table S1.6:** Desethylamodiaquine PK/PD parameters. EC50=half-maximal effective concentration.

| Parameter | Value | Reference |
| --- | --- | --- |
| Absorption rate (per day) | 2·16 | <sup>26</sup> |
| Elimination rate (per day) | 0·86 | <sup>26</sup> |
| Rate at which the drugs move from the central compartment to the peripheral compartment (per day) | 1·70 | <sup>26</sup> |
| Rate at which the drugs move from the peripheral compartment to the central compartment (per day) | 0·46 | <sup>26</sup> |
| Volume of distribution (l/kg) | 18·40 | <sup>26</sup> |
| Maximum killing rate (per day) | 2·30 | <sup>26</sup> |
| EC50 (mg/l) | 0·02 | <sup>26</sup> |
| Slope of the effect curve | 7·00 | <sup>26</sup> |

**Figure S1.7:**
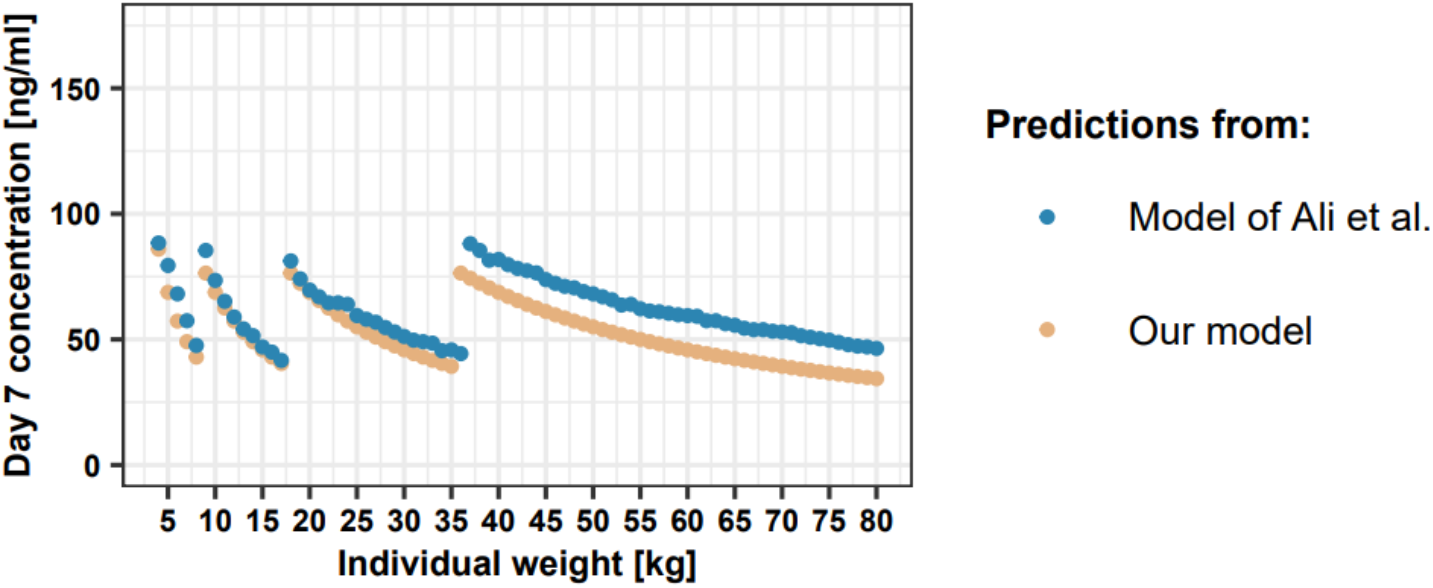
Comparison between the day seven DEAQ concentrations predicted by Ali *et al.*^27^ and our model stratified by child weight. Median day seven DEAQ concentration predicted by Ali *et al.*^27^ (blue dots) and day seven DEAQ concentration predicted by our model (orange dots) stratified by child weight. Predictions from the model of Ali *et al.*^27^ were extracted from Figure 4A of Ali *et al.*^27^, which also shows interquartile ranges that are not displayed here. Ali *et al.* used a dosage regimen based on weight (see **Ali *et al.*** Table 4). To replicate this figure, we used the same dosage regimen as Ali *et al..*^27^ However, for the remainder of this study, the dose regimen was as reported in Table S 1.1.

**Figure S1.8:**
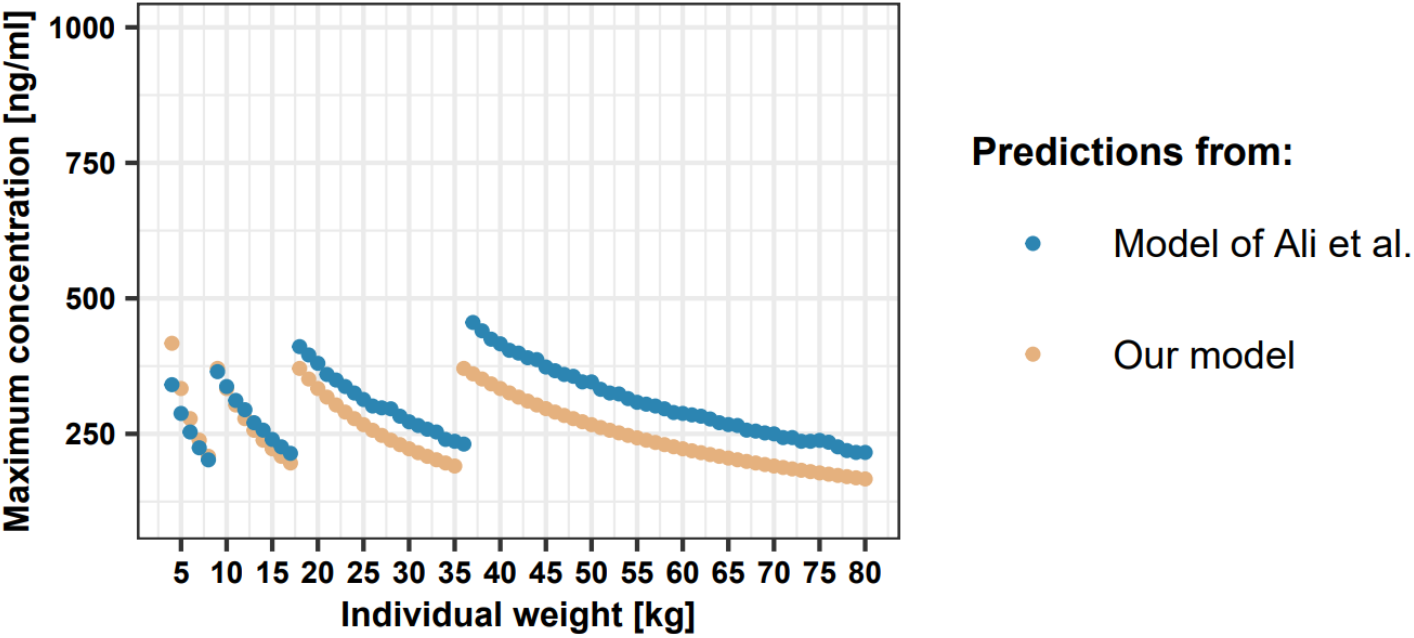
Comparison between the maximum DEAQ concentrations predicted by Ali *et al.*^27^ and our model stratified by child weight. Median maximum DEAQ concentrations predicted by Ali *et al.*^27^ (blue dots) and maximum DEAQ concentrations predicted by our model (orange dots) stratified by child weight. Predictions from the model of Ali *et al.*^27^ were extracted from Figure 4C of Ali *et al.*^27^, which also shows interquartile ranges that are not displayed here. Ali *et al.* used a dosage regimen based on weight (see **Ali *et al.*** Table 4). To replicate this figure, we used the same dosage regimen as Ali *et al..*^27^ However, for the remainder of this study, the dose regimen was as reported in Table S 1.1.

##### 3.3 Replication of the trial of Zongo *et al.*^18^

We compared our parameterisation of SMC with SP*+AQ with results from a randomised non-inferiority trial comparing dihydroartemisinin-piperaquine with SP+AQ.^18^ This trial was conducted in Burkina Faso in 2009. During the trial, three cycles of SMC were deployed to children aged three months to five years during the short transmission season. Children were assigned to either the SP+AQ, dihydroartemisinin-piperaquine, or control group. We parameterised our model to replicate this trial (table S1.6) as was previously done by Burgert *et al.*^28^. We parameterised SP* and AQ as described in Tables S1.3-1.5. In the trial, children who received SP+AQ were mainly infected by the SP-resistant quadruple mutant (41·5% of malaria cases) and more sensitive genotypes (sensitive genotype, single mutant, double mutant, and triple mutant).^18^ Thus, we simulated settings where the parasite population was composed of quadruple mutants (against which SP confers a prophylactic period of 35 days) or of sensitive parasites (against which SP confers a prophylactic period of 42 days). We expected the trial result to lie in between the simulated estimates in these two settings.

Our simulated parasitemia prevalence values for malaria infection in targeted children at the beginning and end of the trial were quite similar to those from Zongo *et al.*^18^ (table S1.7). We also estimated the protective efficacy of SMC against clinical malaria (the relative reduction in the number of clinical malaria cases) over time measured at the last cycle of SMC as was done by Zongo *et al.*^18^ (figure S1.6). As expected, the estimate from Zongo *et al.*^18^ lies in between our estimates when we simulate the trial in a population composed of only quadruple mutants or only of sensitive genotypes. Note that we did not obtain a protective efficacy of 100% in the first days after SMC deployment as was reported by Zongo *et al.*^18^.

This is because in our simulation we assumed children with clinical malaria received artemether-lumefantrine (AL) but not SP+AQ at each SMC deployment. These children receiving AL could be infected sooner than if they had received SP+AQ during SMC, which causes a reduction in SMC protective efficacy at the beginning of the simulation. This factor can also explain the slight increase in efficacy observed in the few days following SMC deployment. For our simulation, the protective effectiveness (PE) does not start to decline until after approximately 20 days with a relatively quick decrease. However, in Zongo et al., the decay of PE is more progressive and starts sooner. This difference may be due to the fact that in the real-world clinical trial, there is more variation in exposure and PP between individuals than assumed in our model due to the variation of PK parameters between individuals (see discussion). These results suggest that our parameterisation could accurately recover the protective efficacy of SP+AQ against clinical malaria observed by Zongo *et al.*^18^ even if we parameterised SP*’s prophylactic period to match clinical data on malaria infections.

**Table S1.7:** Model inputs to replicate the study by Zongo *et al.*^18^. The level of access to treatment was estimated to reproduce the parasitemia prevalence in children targeted at the beginning of the trial of Zongo et al.^18^ (see table S1.7). We parameterised SP* and AQ as described in Tables S1.3-5. AL=artemether-lumefantrine. AQ amodiaquine. EIR=entomological inoculation rate. ITN=insecticide-treated net. Ref= reference. SP=sulfadoxine-pyrimethamine.

| Model component | Parameter | Value | Ref. |
| --- | --- | --- | --- |
| Vector specification | Mosquitos species (% of total abundance) | *A. funestus*: 42% indoor, 31% outdoor<br>*A. gambiae*: 14% indoor, 13% outdoor | 28,29 |
|  | Annual EIR (inoculations per person per year) | 160 | 28,29 |
|  | Seasonality of malaria transmission (inoculations per person per year) | *A. funestus*: August: 26, September: 16, October: 0·3, November: 1<br>*A. gambiae*: August: 40, September: 50, October: 16, November: 6·4 | 28,29 |
| Heath system | Probability that symptomatic cases receive treatment within two weeks from symptom onset (%). The first-line treatment was AL. | 2003-2010: 20% | Estimated |
| Vector interventions | Coverage of ITN | 2006-2008: 14%<br>2009-2010: 27% | 18,28,30 |
| SP+AQ group | Deployment timing (children with fever history were tested for malaria: if negative: children received SP+AQ; if positive: children received AL) | 1<sup>st</sup> cycle : 15 August, 2<sup>nd</sup> cycle : 15 September, 3<sup>rd</sup> cycle : 15 October | 18 |
|  | Follow up (children with fever history were tested for malaria: if positive: children received AL) | 1<sup>st</sup> cycle : 1 September, 2<sup>nd</sup> cycle : 1 October, 3<sup>rd</sup> cycle : 1 November | 18 |
| Control group | Deployment timing (children with fever history were tested for malaria: if positive: children received AL) | 1<sup>st</sup> cycle : 15 September, 2<sup>nd</sup> cycle : 15 October | 18 |
|  | Follow up (children with a history of fever were tested for malaria: if positive: children received AL) | 1<sup>st</sup> cycle : 1 October, 2<sup>nd</sup> cycle : 1 November | 18 |

**Table S1.8:**
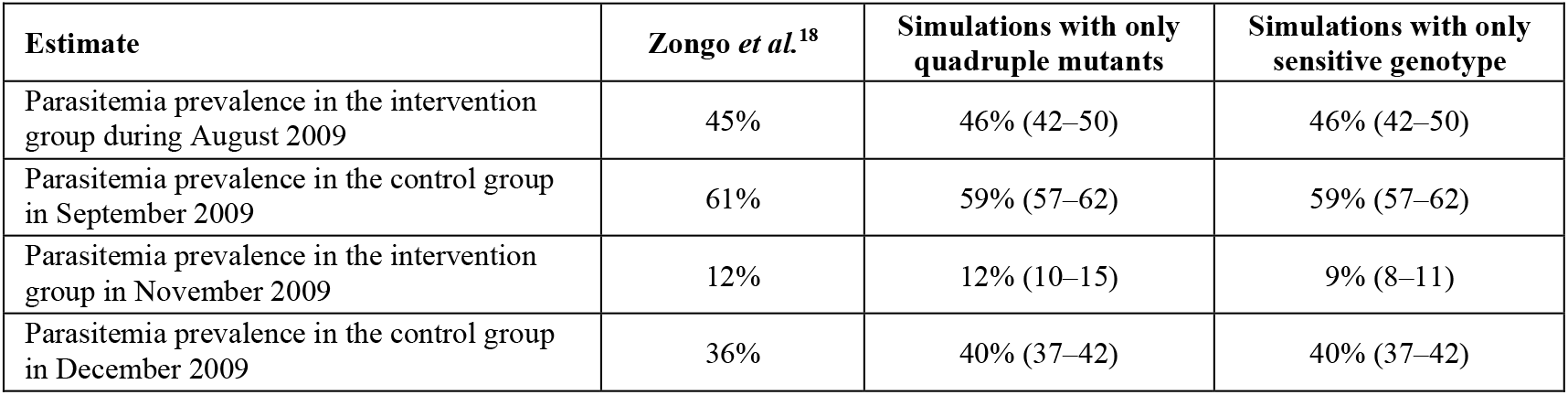
Comparison between the parasitemia prevalence estimated at different time points in the trial by Zongo *et al.* ^18^ and from our model. In the study of Zongo *et al.* prevalence values were estimated for children aged three months to five years in August and September 2009 (i.e. prior to SMC) and in November and December 2009 (after SMC), see table S1.6 for details. **We assumed differences between prevalence in August and September (prior to SMC implementation in the intervention group) were due to the increase in transmission intensity.** Our model simulated 20 stochastic realisations of the trial with the parasite population composed of either the quadruple mutant or the SP-sensitive parasite. The 95% confidence intervals are shown in parentheses. The prevalence assuming quadruple or sensitive genotypes was the same in August and September as we assumed no SP was used prior to SMC. The only difference in prevalence between the quadruple mutant and sensitive genotype occurred in November after SMC implementation. As SMC was more efficient against the sensitive genotype, prevalence was lower in populations of sensitive genotypes compared to populations of the quadruple mutant. In December 2009 of the study, the prevalence of assuming only quadruple or sensitive genotypes in the population was the same in the control group, as no SP was used in the control group.

**Figure S1.9:**
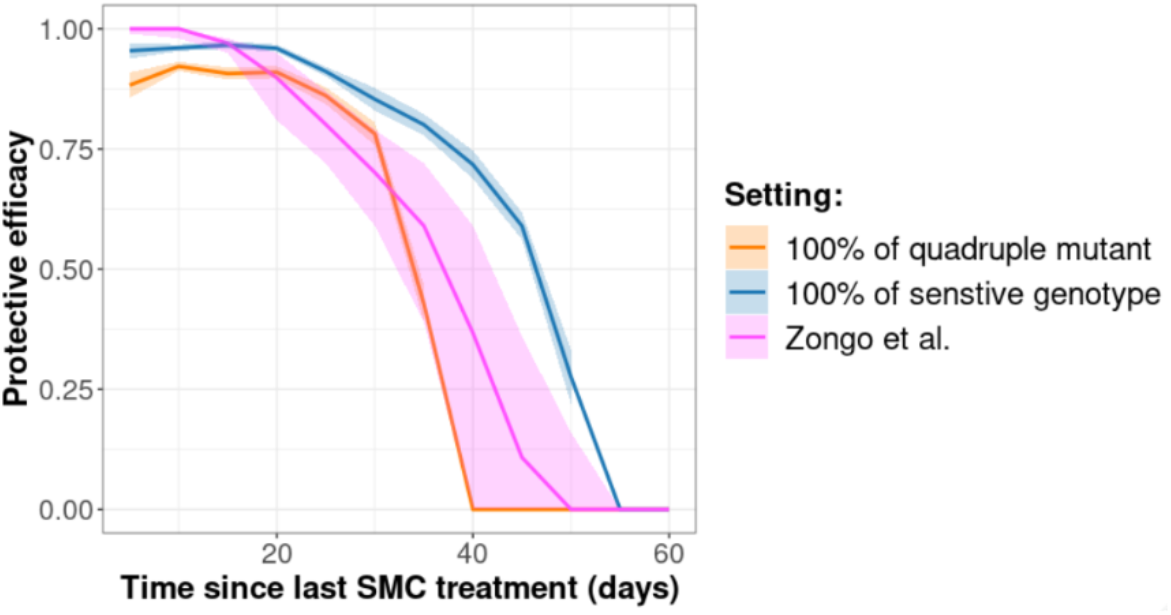
Protective efficacy decay of SMC with SP+AQ estimated by Zongo *et al.*^18^ and from our model. The decay of the protective efficacy against clinical malaria was estimated at the last cycle of SMC. The purple curve represents the protective efficacy estimated by Zongo et al.^18^ (extracted from Figure 3 of Zongo et al.^18^). The orange and blue curves highlight the mean protective efficacy estimated over 20 stochastic simulations when we assumed that the parasite population was composed of either quadruple mutants or SP-sensitive genotypes, respectively. Shaded areas indicate 95% confidence intervals. The fact that simulation start at PE less that 1.0 is explained in section 1.2.3 above.

### 4 Parameterisation of first-line treatment

The first-line treatment was assumed to be an artemisinin-based combination therapy (ACT). We modelled dihydroartemisinin as the artemisinin derivative (table S1.8). To ensure that the long-acting drug had the profile of typical partner drugs, we used the PK/PD parameter of piperaquine (table S1.9), but we varied the elimination rate to capture the impact of the partner drug half-life on the rate of spread of the quintuple mutant in the global sensitivity analysis. The elimination rate range in table S1.9 allowed us to capture the half-lives of lumefantrine, piperaquine, and mefloquine (Table 1). The ACT was fully and clinically equally efficient against quadruple and quintuple mutants. The dosage was a daily dose of dihydroartemisinin (4 mg/kg) and the partner drug (30 mg/kg) for three days.

**Table S1.8:**
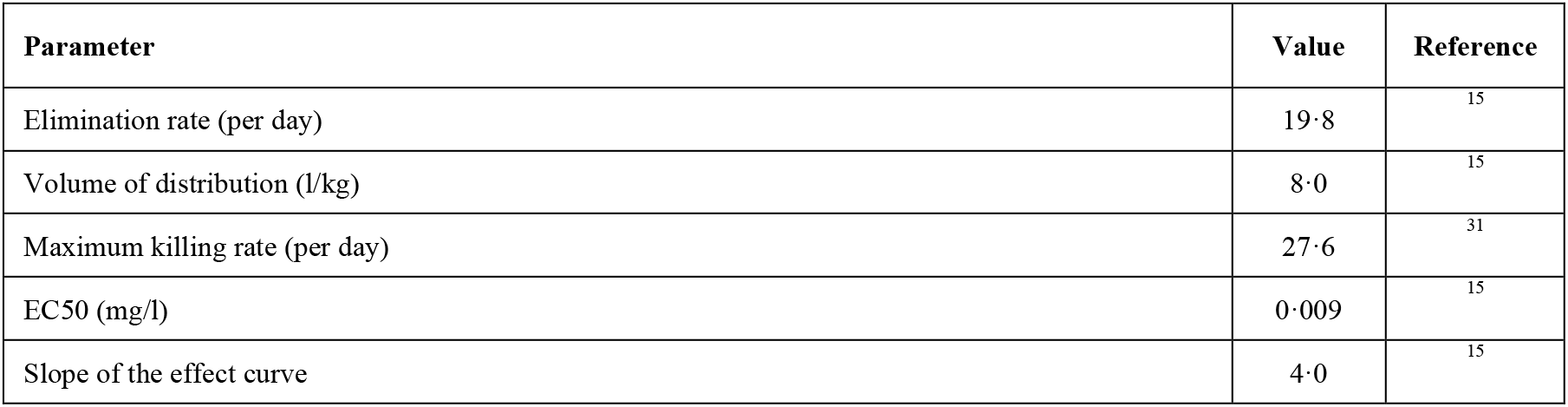
Dihydroartemisinin PK/PD parameters. EC50=half-maximal effective concentration.

| Parameter | Value | Reference |
| --- | --- | --- |
| Elimination rate (per day) | 19·8 | <sup>15</sup> |
| Volume of distribution (l/kg) | 8·0 | <sup>15</sup> |
| Maximum killing rate (per day) | 27·6 | <sup>31</sup> |
| EC50 (mg/l) | 0·009 | <sup>15</sup> |
| Slope of the effect curve | 4·0 | <sup>15</sup> |

**Table S1.9:** Partner drug’s PK/PD parameters. EC50=half-maximal effective concentration.

| Parameter | Value | Reference |
| --- | --- | --- |
| Absorption rate (per day) | 11·16 | <sup>32</sup> |
| Rate at which the drug moves from the central compartment to the peripheral compartment (per day) | 8·46 | <sup>32</sup> |
| Rate at which the drug moves from the peripheral compartment to the central compartment (per day) | 3·30 | <sup>32</sup> |
| Elimination rate (per day) | 0·31-1·17 | <sup>32</sup> |
| Volume of distribution (l/kg) | 173·0 | <sup>32</sup> |
| Maximum killing rate (per day) | 3·45 | <sup>15</sup> |
| EC50 (mg/l) | 0·03 | <sup>15</sup> |
| Slope of the effect curve | 6·0 | <sup>15</sup> |

### 5 Supplementary methods and results

#### 6 Assessment of the impact of factors on the rate of spread

We systematically assessed the impact of multiple factors (see main text Table 1) on the spread of the quintuple mutant through global sensitivity analyses. Each global sensitivity analysis required many simulations (200,000 simulations; see details below). It was computationally unfeasible to run such a large number of simulations in OpenMalaria for each global sensitivity analysis. To be computationally efficient, we used a previous approach we developed to assess the impact of factors on the spread of parasites resistant to first-line treatment.^4^ This approach involved training an emulator on a set of OpenMalaria simulations (2,500 simulations; see details below). Emulators are predictive models that can approximate the relationship between input and output parameters of complex models and run much faster than complex models. Then, we performed the global sensitivity analyses on trained emulators. The steps of the approach are described in figure S2.1 and summarised below.

##### i. Randomly sample combinations of parameters

First, we randomly sampled 250 parameter combinations from the parameter ranges (see main text Table 1) using a Latin hypercube sampling algorithm (LHS).^33^ The parameter ranges were defined as follows. The range of SMC coverage corresponded to the range reported by an implementation study.^34^ The variation in EIR captured settings with low transmission to those with high transmission. The range of access to treatment captured low to high levels of access to treatment. The first-line treatment was an artemisinin-based combination therapy (ACT) using a partner drug other than SP or AQ, as recommended by the WHO.^16^ Children in the targeted age groups could still contract malaria infections and obtain first-line treatment through the formal health sector similarly to individuals not targeted by SMC. The ACT was effective against both quintuple and quadruple infections. We varied the half-life of the long-acting partner drug in a range that captured the half-life of lumefantrine, mefloquine, and piperaquine.^26, 35–38^

##### ii. Model simulation and estimation of selection coefficients

For each parameter combination, we simulated and quantified the rate of spread of the quintuple mutant in our model as follows. Each parameter combination was simulated in five seeds and modelled a human population of 100,000 individuals with demographic age distribution of Tanzania, a lower-middle-income country.^39^ Simulations started with a burn-in period of 130 years. After the burn-in period, we deployed SMC for ten consecutive years and estimated the spread of the quintuple mutant through the selection coefficient which measures the rate at which the logit of the resistant genotype frequency increases each parasite generation (see below).^40^ Note that simulations started with a frequency of the quintuple mutant in infection of 50%. We did not start with a lower frequency because the strength of genetic drift would have been more substantial, which would have led to stochastic extinction of the quintuple mutant in many simulations and reduced the accuracy of the estimated selection coefficient.^4, 40^ Nevertheless, as the selection coefficient is not frequency-dependent in our model, our estimates represent selection that occurs at a low frequency of the quintuple mutant.^4, 40^

To calculate the selection coefficient, we defined the frequency of the quintuple genotype, *p*, based on the frequency of the quintuple mutant in inoculations (the number of inoculations carrying the resistant genotype divided by the sum of the number of inoculations carrying the resistant genotype and the number of inoculations carrying the sensitive genotype, figure S2.2). This measurement is a good representation of the spread at the population level as it tracked genotypes transmitted to the next generations and was more stable across age groups than other variables, such as the number of patients infected by each genotype (figure S2.3). The selection coefficient, *s,* was defined as the slope of the logistic regression of *p*(*t*),

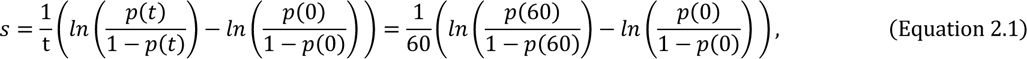

where *t* is the number of parasite generations after the deployment of SMC at *t* = 0. We assumed that a parasite generation is equal to 60 days (6 generations per year).^40^ We started the regression directly at the first cycle of SMC (*t* = 0), and we stopped the regression ten years later at the last SMC cycle (t = 60). We stopped the regression sooner if *p* was higher than 90% or lower than 30% to prevent tracking one genotype at a low frequency which could have led to strong genetic drift.

##### iii. Model emulation and adaptive sampling

Once the selection coefficient was estimated for each parameter combination, we randomly split our data into a training and a testing dataset containing 80% and 20% of the simulations, respectively. Then, we trained a heteroskedastic Gaussian process (HGP) on the training dataset using the function *mleHetGP* from the R-package *hetGP*.^41^ We chose to use HGP as it was successfully used in previous studies that performed global sensitivity analysis of OpenMalaria.^4, 11, 42^ To assess the accuracy of the emulator, for the test dataset, we assessed the correlation coefficient and root mean squared error between selection coefficients predicted with the emulator and selection coefficients estimated using OpenMalaria. If the fit of the emulator was not satisfactory, we iteratively improved the emulator fit through adaptive sampling. To undertake adaptive sampling, we sampled 100 parameter combinations in the parameter space region where we were less confident (higher variance) in the emulator prediction, and we repeated the whole process until the emulators reached sufficient accuracy (figure S2.4).

##### iv. Global sensitivity analysis

After emulation and adaptive sampling, we assessed the influence of each factor on the selection coefficient via global sensitivity analysis using Sobol’s method of variance decomposition.^43^ To perform the global sensitivity analysis, we first generated two random datasets of 100’000 parameter combinations using Latin hypercube sampling. Then, we predicted the selection coefficient for each parameter combination using the emulators. Note that without emulators, we would have to run these 200’000 simulations in OpenMalaria for each global sensitivity analysis, which would not have been feasible due to computational requirements. Finally, we performed the global sensitivity analysis on the two datasets with 150000 bootstrap replicates using the function *soboljansen* of the R-package *Sensitivity*.^44^ With the global sensitivity analysis, we estimated the first-order indices of each factor, representing their influence on the rate of spread (figure S2.5), and assessed the 25th, 50th, and 75th quantiles of the predicted rate of spread of the two random datasets over each parameter range (figure S2.6).

**Figure S2.1:**
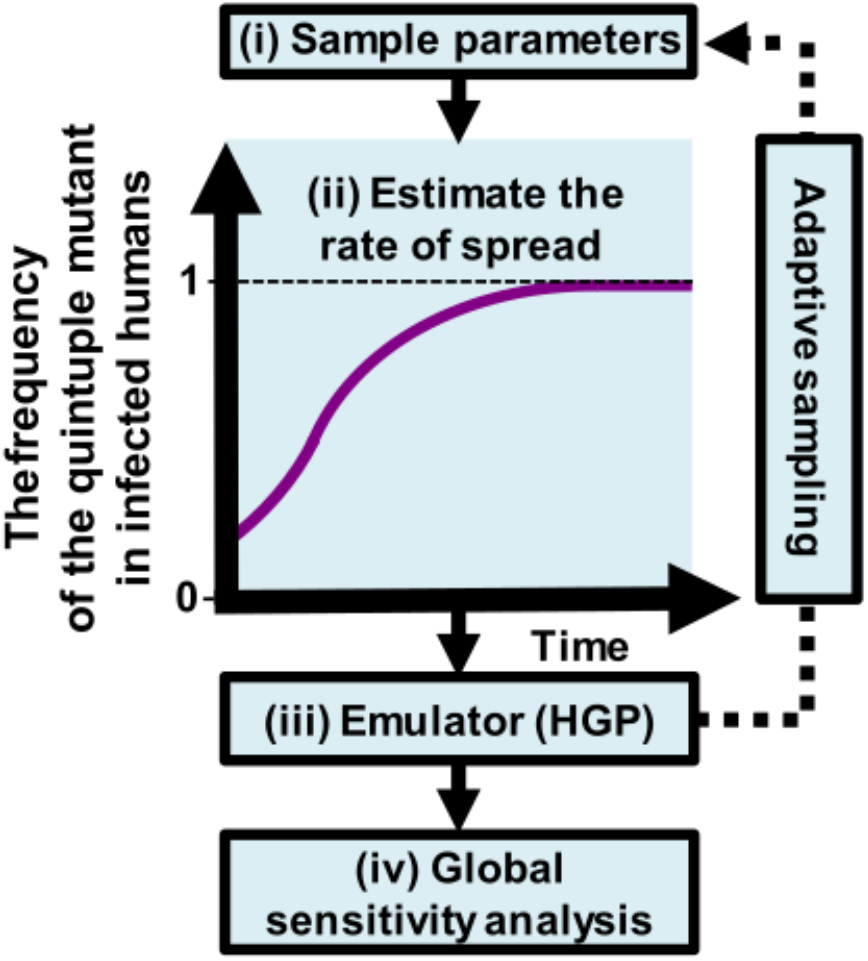
Schematic of our global sensitivity analysis approach. *The approach aims to assess the influence of factors on the rate of spread (selection coefficient) of the quintuple mutant through global sensitivity analyses. Our approach involved: (i) randomly sampling parameters combinations; (ii) simulating and estimating the rate of spread of the quintuple mutant via the selection coefficient for each parameter combination with our model; (iii) training an emulator on the simulated data with iterative fitting improvements through adaptive sampling, and (iv) performing the global sensitivity analysis using the fitted emulator. The purple line illustrates an example of the frequency of the quintuple mutant in infected humans across time. HGP:* Heteroskedastic Gaussian Process.

**Figure S2.2:**
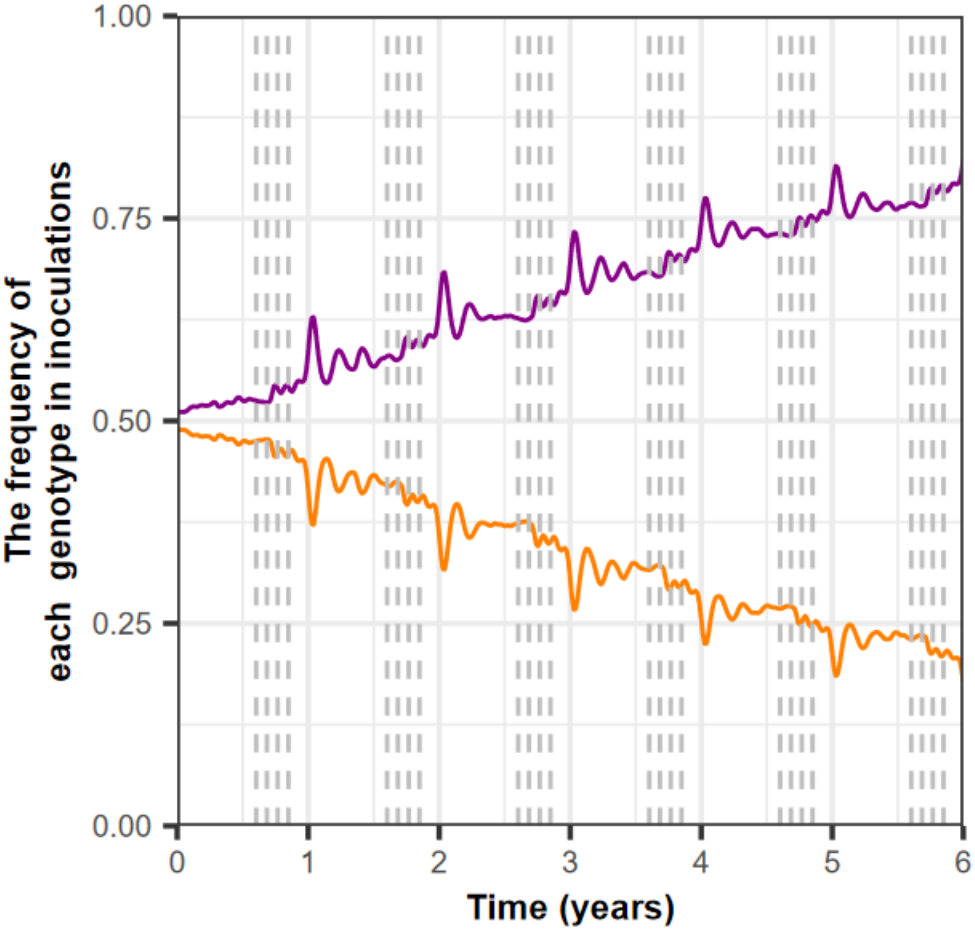
The frequency of each genotype in inoculations. We estimated the frequency of inoculations carrying the quadruple (orange curve) and quintuple mutants (purple curve) when we deployed SMC four times per year in the moderate seasonal setting to children aged three months to ten years. The coverage was equal to 98% at each cycle. The EIR was equal to 339 inoculations per person per year. The access to treatment was equal to 35%. The grey lines indicate SMC deployment.

**Figure S2.3:**
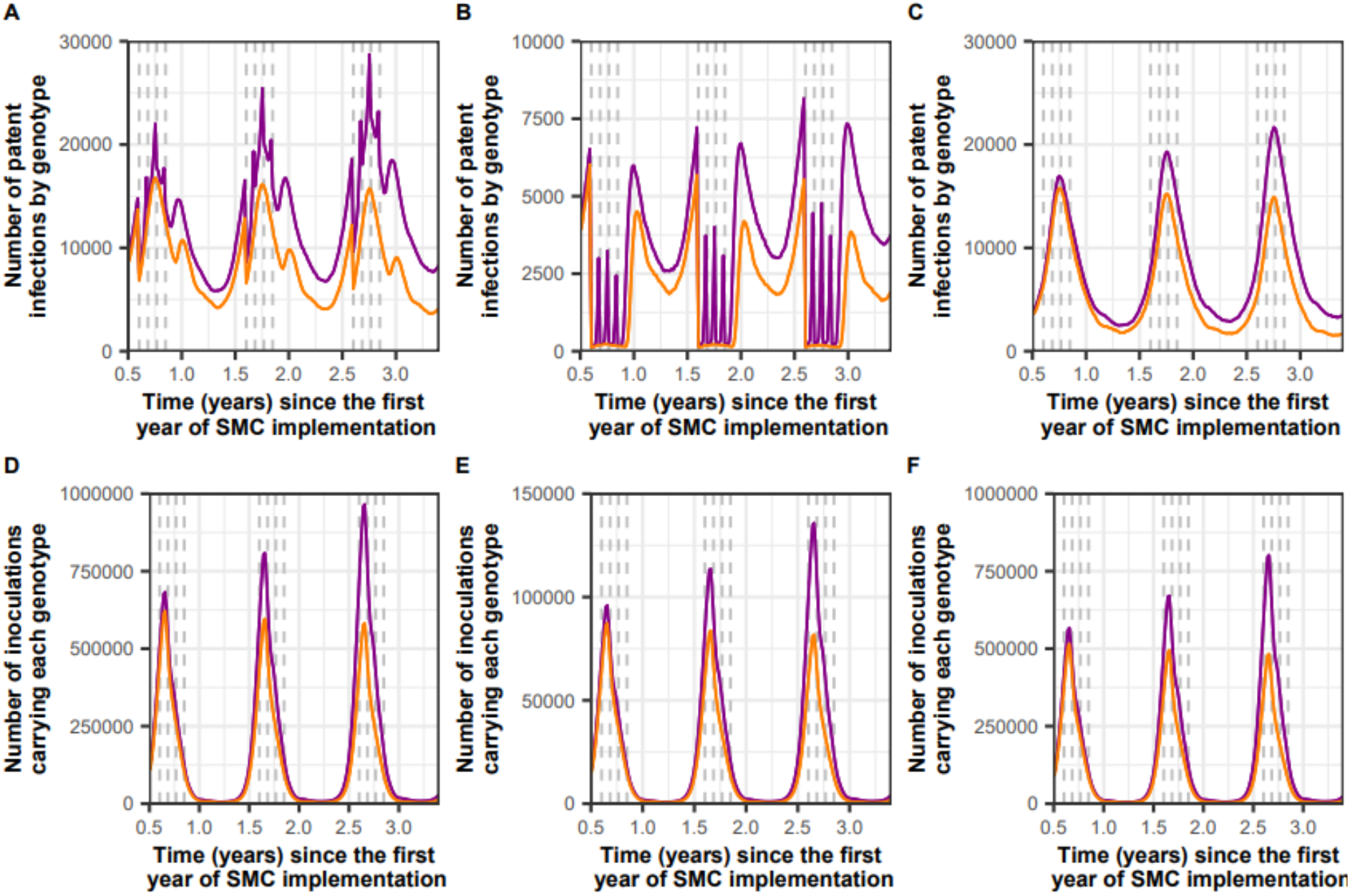
Illustration of the number of the quadruple and quintuple mutants in infections and inoculations in different age groups. The top row shows the number of patent infections caused by the quadruple mutant (orange lines) and the quintuple mutant (purple lines) in (A) the total population, (B) children under ten years old, and (C) individuals over ten years old. The bottom row shows the number of inoculations caused by the quadruple mutant (orange lines) and the quintuple mutant (purple lines) in (D) the whole population, (E) children under ten years old, and (F) individuals over ten years old. In each plot, we delivered SMC four times a year (grey dashed lines) in the moderate seasonal setting to children under ten years with a coverage of 98% (high coverage). The EIR was equal to 339 inoculations per person per year (high transmission). The access to treatment was equal to 35% (low access).

**Figure S2.4:**
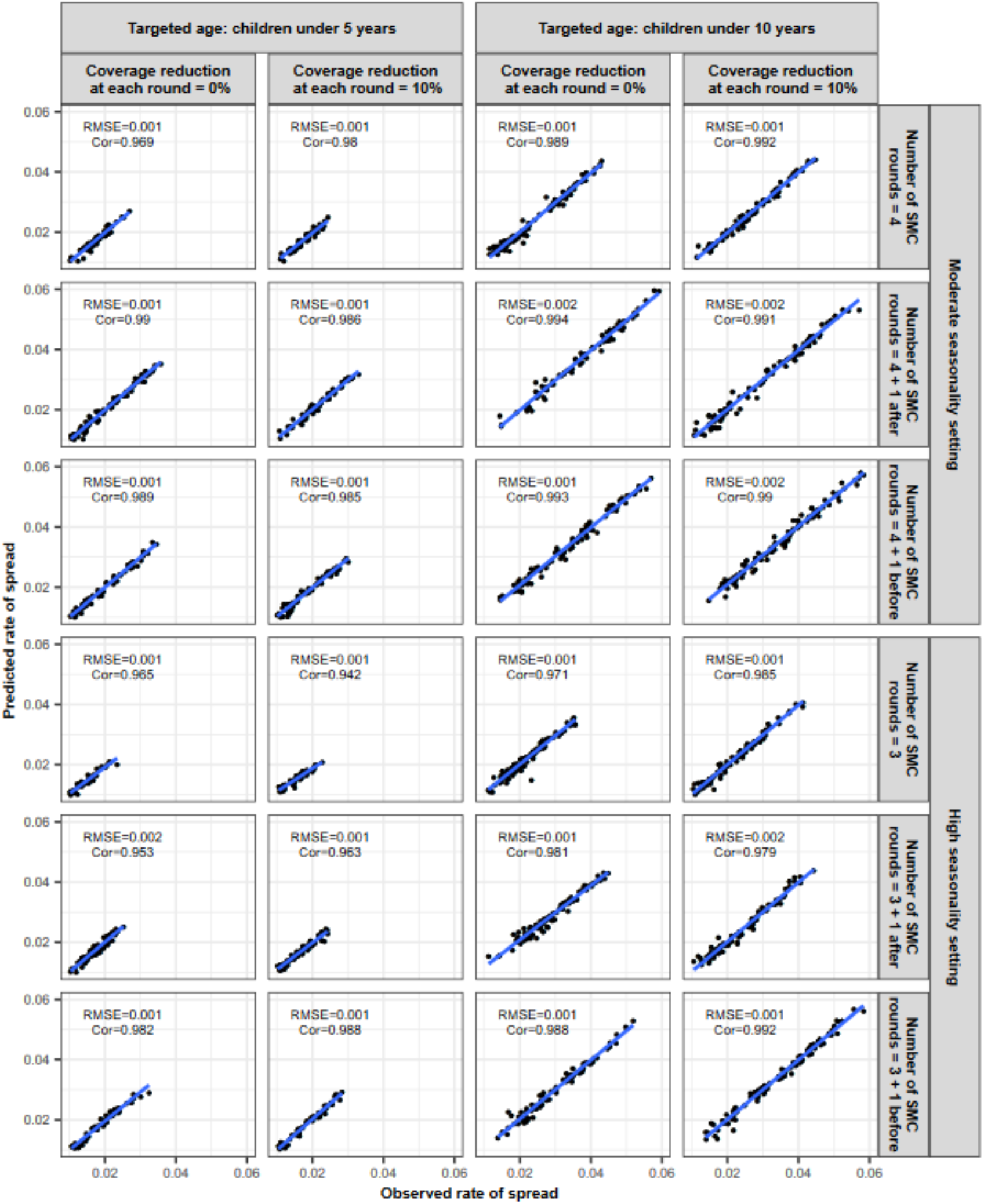
Accuracy of the emulators used for each global sensitivity analysis of the spread of the quintuple mutant in each seasonality setting and for each SMC deployment strategy. The figure exhibits the observed rate of spread of the quintuple mutant estimated by our model simulations and the rate of spread predicted with the emulators for the testing dataset at the final cycle of adaptive sampling for each global sensitivity analysis. We performed the global sensitivity analyses in two different seasonality settings (moderate and high seasonal) and for various SMC deployment strategies that varied by: targeted age groups (children aged three months to five years or children aged three months to ten years), numbers of SMC cycles deployed per year (high seasonality setting: three cycles or four cycles with the additional cycle deployed before or after the typically implemented deployment regime of SMC, moderate seasonality setting: four cycles or five cycles with the additional cycle deployed before or after typically implemented deployment regime of SMC), and assumptions about the reduction of the coverage across each cycle (none (0%) or 10% from the previous cycle). Cor=Spearman correlation coefficient, RMSE=root means squared error, blue lines: linear regression fit.

**Figure S2.5:**
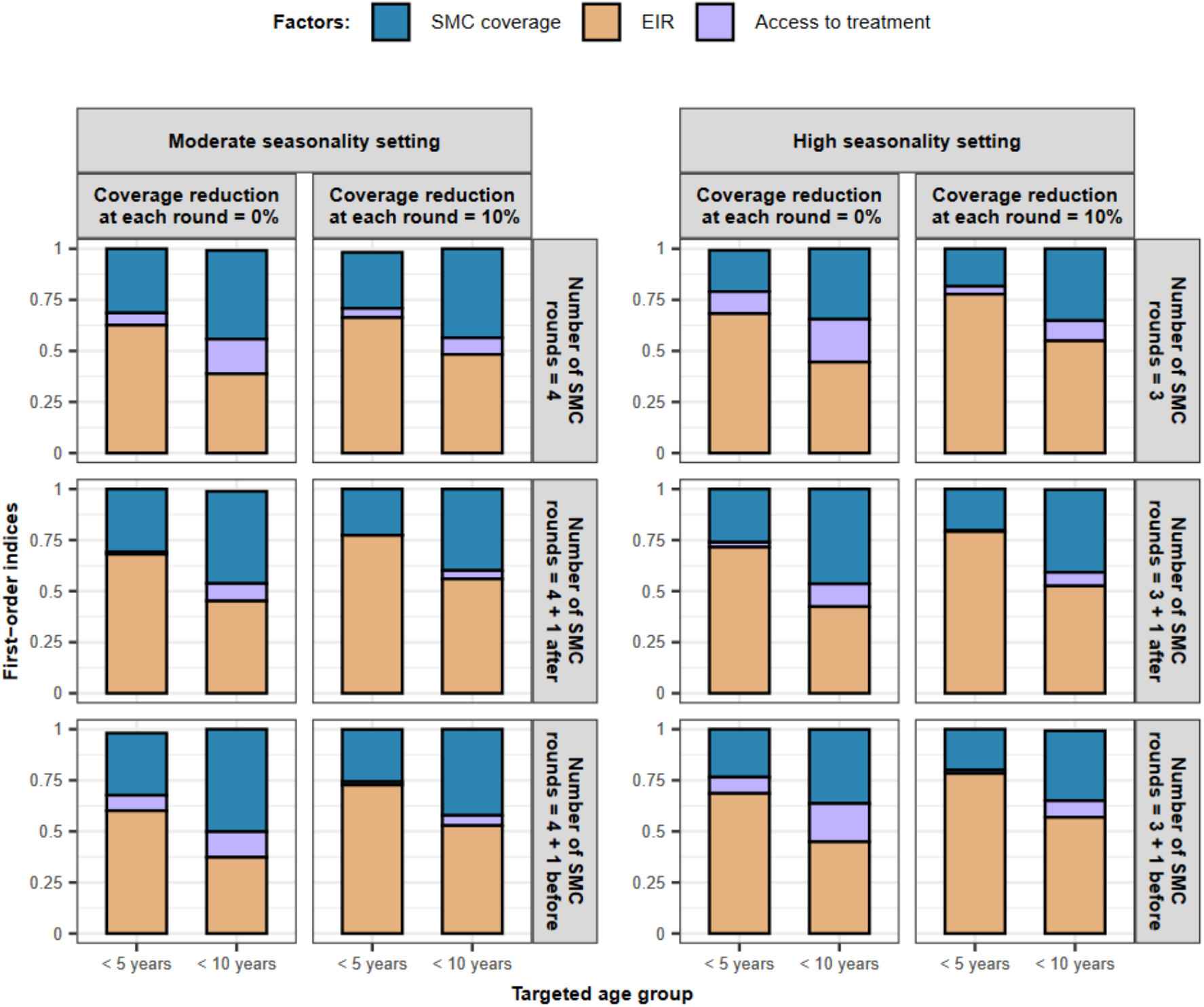
The impact of factors on the spread of the quintuple mutant. The figure displays the first-order indices of the coverage (blue), the EIR (orange), and the level of access to treatment (purple) estimated during each global sensitivity analysis. In the figure, the first-order indices of each factor are proportional to their size within the rectangle and represent their influence on the rate of spread. The explored parameter ranges were the following: SMC coverage [70%, 100%]; EIR [5, 500] inoculations per person per year; and the level of access to treatment [10%, 80%]. We performed the global sensitivity analyses in two different seasonality settings (moderate and high seasonal) and for various SMC deployment strategies that varied by: targeted age groups (children aged three months to five years or children aged three months to ten years), numbers of SMC cycles deployed per year (high seasonality setting: three cycles or four cycles with the additional cycle deployed before or after the typically implemented deployment regime of SMC, moderate seasonality setting: four cycles or five cycles with the additional cycle deployed before or after the typically implemented deployment regime of SMC), and assumptions about the reduction of the coverage across each cycle (none (0%) or 10% from the previous cycle).

**Figure S2.6:**
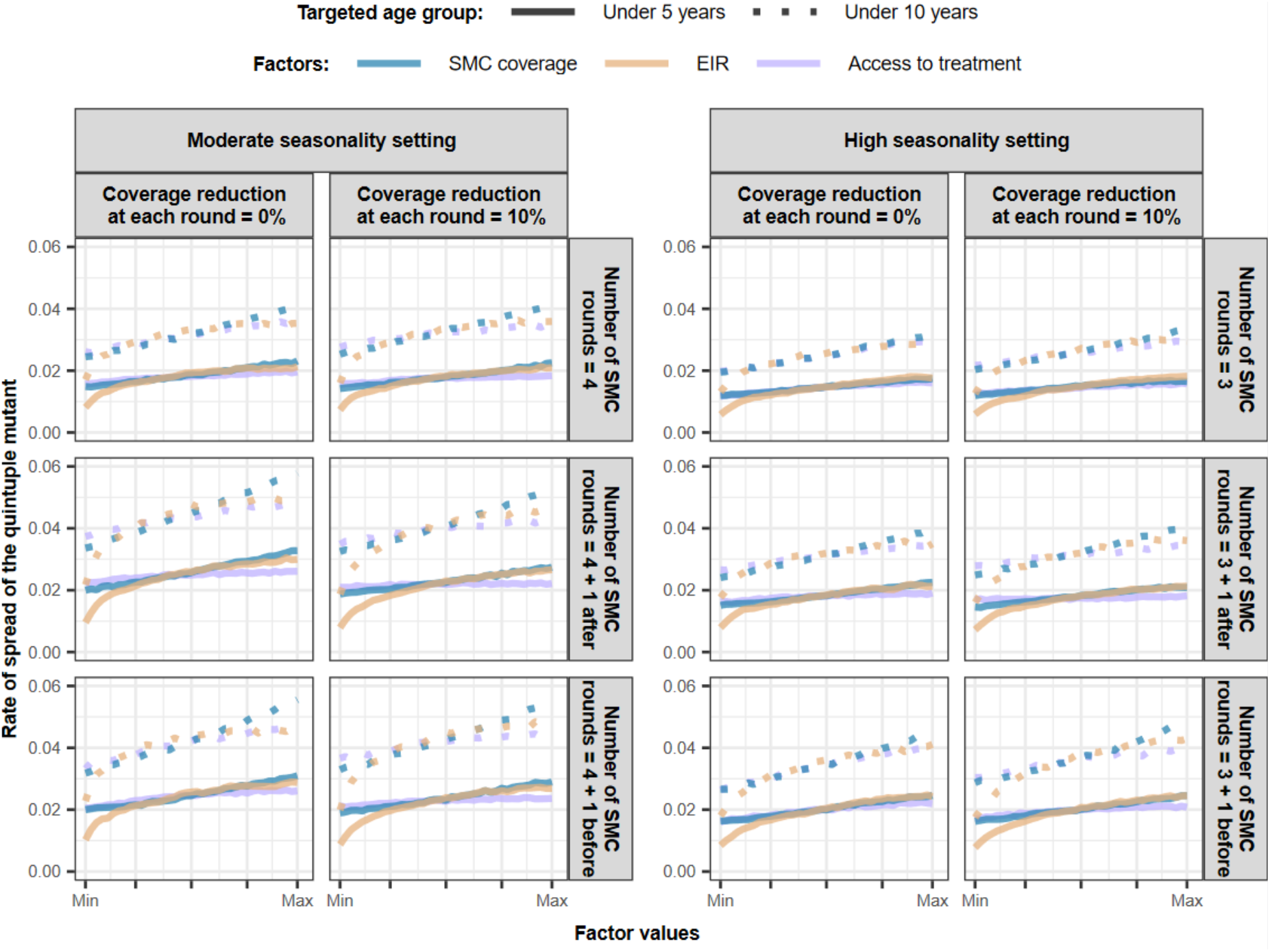
Influence of different factors on the rate of spread of SP-resistant quintuple mutant. *Curves represent the predicted median rate of spread (selection coefficient) of the quintuple mutant estimated from the global sensitivity analysis over the key parameter ranges for each epidemiological, setting specific, SMC deployment strategy, or drug property factor when SMC is delivered to children between three months and five years of age (solid lines) or children between three months and ten years (dashed lines). The factors and their parameter ranges were as follows: SMC coverage at cycle one during the transmission season [70%, 100%] (blue lines); transmission level represented by EIR [5, 500] inoculations per person per year (orange lines); and the level of access to treatment [10%, 80%] (purple lines). The half-life of the partner drug of the first-line ACT *([6, 22] days) is not displayed on the figure as it did not impact the rate of spread. Results are for different seasonality settings (moderate or high seasonality), for various numbers of cycles deployed per year (high seasonality setting: three cycles or four cycles with the additional cycle deployed before or after the* typically implemented *SMC deployment regime; moderate seasonality setting: four cycles or five cycles with the additional cycle deployed before or after the* typically implemented SMC *deployment regime), and different assumptions around the reduction of the SMC coverage across each cycle (0% (constant coverage) or 10% reduction* (e.g., if the first cycle is 80%, then subsequent cycles are 72%, 65%, etc.).* ACT=artemisinin-based combination therapy. EIR=entomological inoculation rate. SMC=seasonal malaria chemoprevention.

**Figure S2.7:**
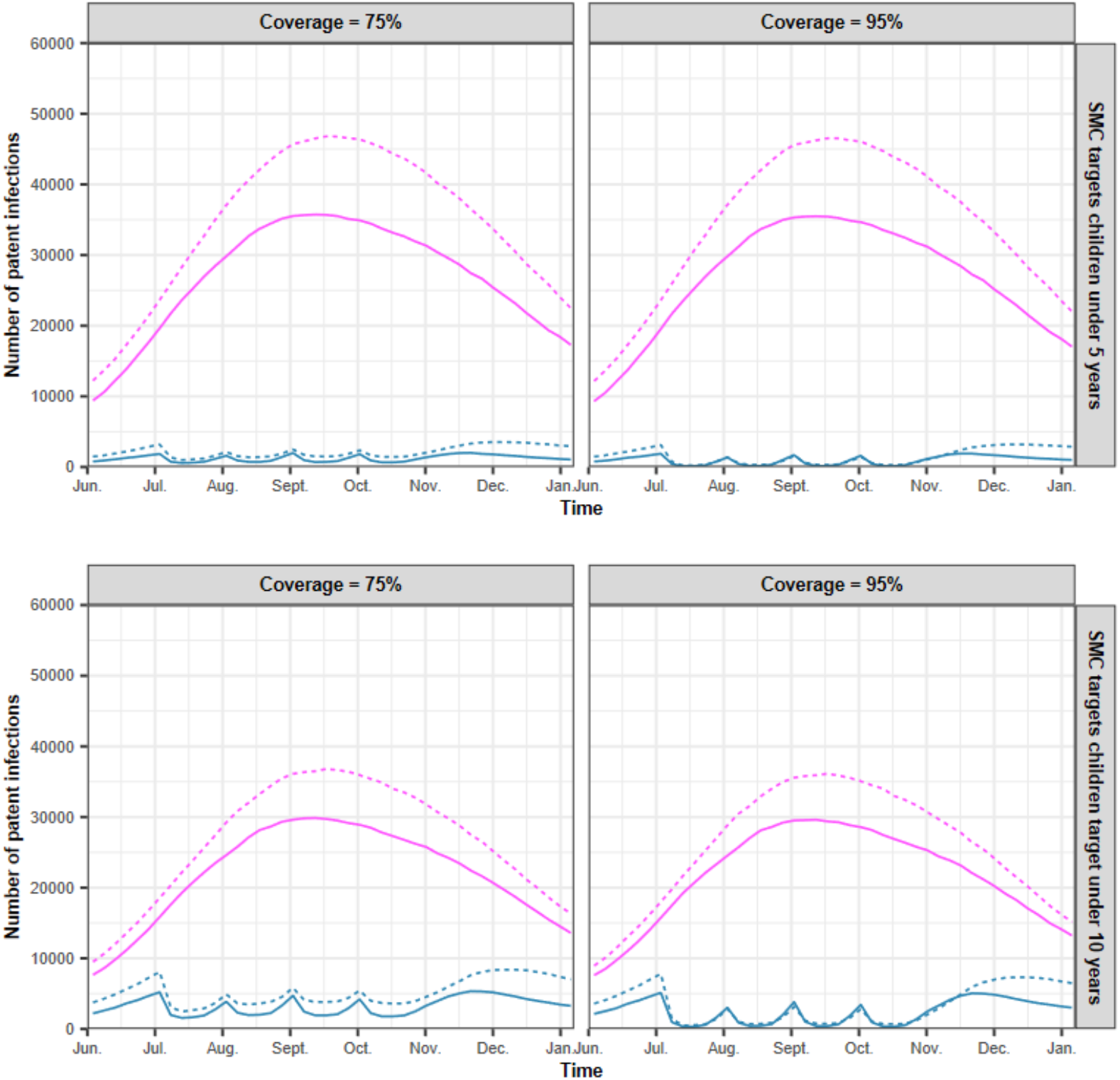
The impact of access to first-line treatment on the number of patent infections. *The figure reports the numbers of patent infections in children targeted by SMC (*three months to *five years old or three months to ten years) (blue curves) and in individuals who were not targeted by SMC (above five or ten years old) (pink curves) in settings where the level of access to treatment was low (solid line, access to treatment: 10%) or high (dashed line, access to treatment:* 80*%). In this example, four cycles of SMC were deployed in the moderate seasonal setting to children* aged three months to *five years or children aged three months to ten years. The coverage was equal to 75% or 95% for each SMC cycle. The coverage was equal to 95% for each SMC cycle. The EIR was equal to 50 inoculations per person per year (high transmission)*.

**Figure S2.8:**
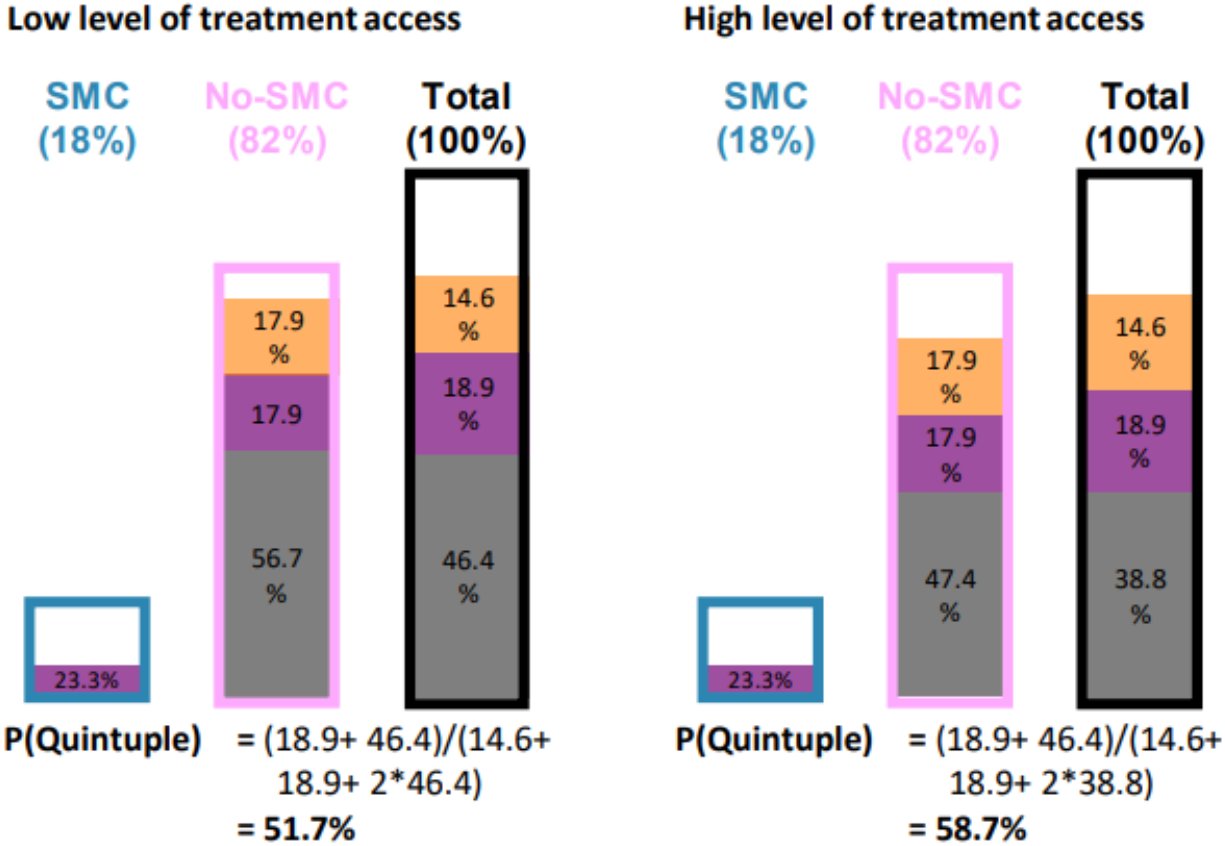
The effect of access to treatment on the frequency of the quintuple mutant. The figures schematically illustrate the effect of access to treatment on the frequency of the quintuple mutant*. The frequencies were estimated when the frequency of each mutant was equal to 50% and at the time step before we deployed a third cycle of SMC to children* aged three months to *five years in settings that had a low (10%) or high (80%) level of access to treatment. The SMC coverage was 95% and constant across cycles. The EIR was equal to 50 inoculations per person per year (high transmission). The percentages of individuals targeted by SMC are in the blue bars, and those not targeted by SMC are in the pink bars. The black bars represent the total population. Within each bar, individuals are separated as carrying no parasites (white), the quadruple mutant (*orange*), the quintuple mutant (purple), or both mutants (grey). The frequency of the quintuple mutant in infected individuals, P(Quintuple), is the number of infections caused by the quintuple mutant to the total number of infections. Note that the level of mixed infection is high in this illustration because we assumed that the diagnostic test could detect all infections (assuming no detection limit for the diagnostic tools)*.

#### 7 Conversion from the selection coefficient into the *T50*

To illustrate our results in a given time frame, we translated the selection coefficient to the time needed for the quintuple mutant to spread from 1% (the current frequency in the Sahel) to 50% frequency of inoculations, *T_50_*, for a set of parameter combinations. We first predicted the selection coefficient with the trained emulators for each deployment strategy for three coverage levels (75%, 85%, and 95%), with transmission ranging from 5 to 150 inoculations per person per year, and with access to treatment of 25% and 70%. Then, we converted the selection coefficients to *T_50_*. To convert the selection coefficient into the *T_50_* we converted the selection coefficient into the numbers of parasite generations needed for the frequency of the quintuple mutant in inoculations to increase from 1% to 50% as follow, *t*,

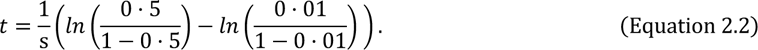

Then, we converted the number of parasite generations to the *T_50_* in years. We assumed that a parasite generation is equal to 60 days.^40^ We also illustrated the *T_50_* for different levels of EIR (figure S2.9) or prevalence (figure S2.10) and when we deployed SMC with an equitable delivery (deployment where each child had the same probability of receiving SMC each cycle) (figure S2.11).

We chose to show the times for the mutation to spread from 1% to 5%, but it is straightforward to convert these to other frequency periods. For example, current frequency may already be relatively high at 5%, and a reader may want to convert the illustrated T_50_ to the time taken to spread from 5% to 50%, which we denote t(5→50). It is simply a case of recalibrating Equation 2.2 to get a conversion factor, f. In the 5% to 50% example, f is calculated as

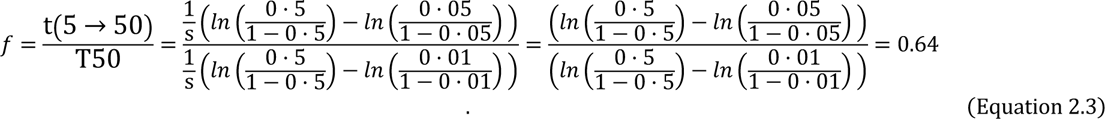

So a typical value of 50 years for T_50_ (e.g. Figure 3 of main text, figure S2.9, S2.10, S2.11 below) would fall to 50*0.64=32 years for t(5→50), a value of 20 years for T_50_ would fall to 20*0.64=12.8 years for t(5→50), and so on.

Similarly, if a reader suspects starting frequency may be low at 0.1% but that SMC efficacy would decline at around 25% frequency, then T50 could be converted to t(0.1→25) using the scaling factor

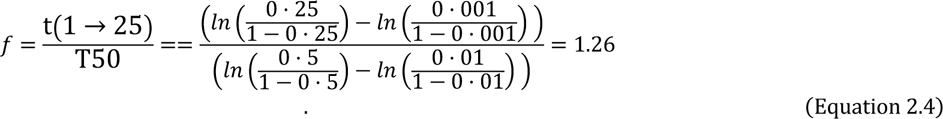

Note that the sensitivity analyses were based on ‘s’ rather than T_50,_ so apply to spread over any frequency periods.

**Figure S2.9:**
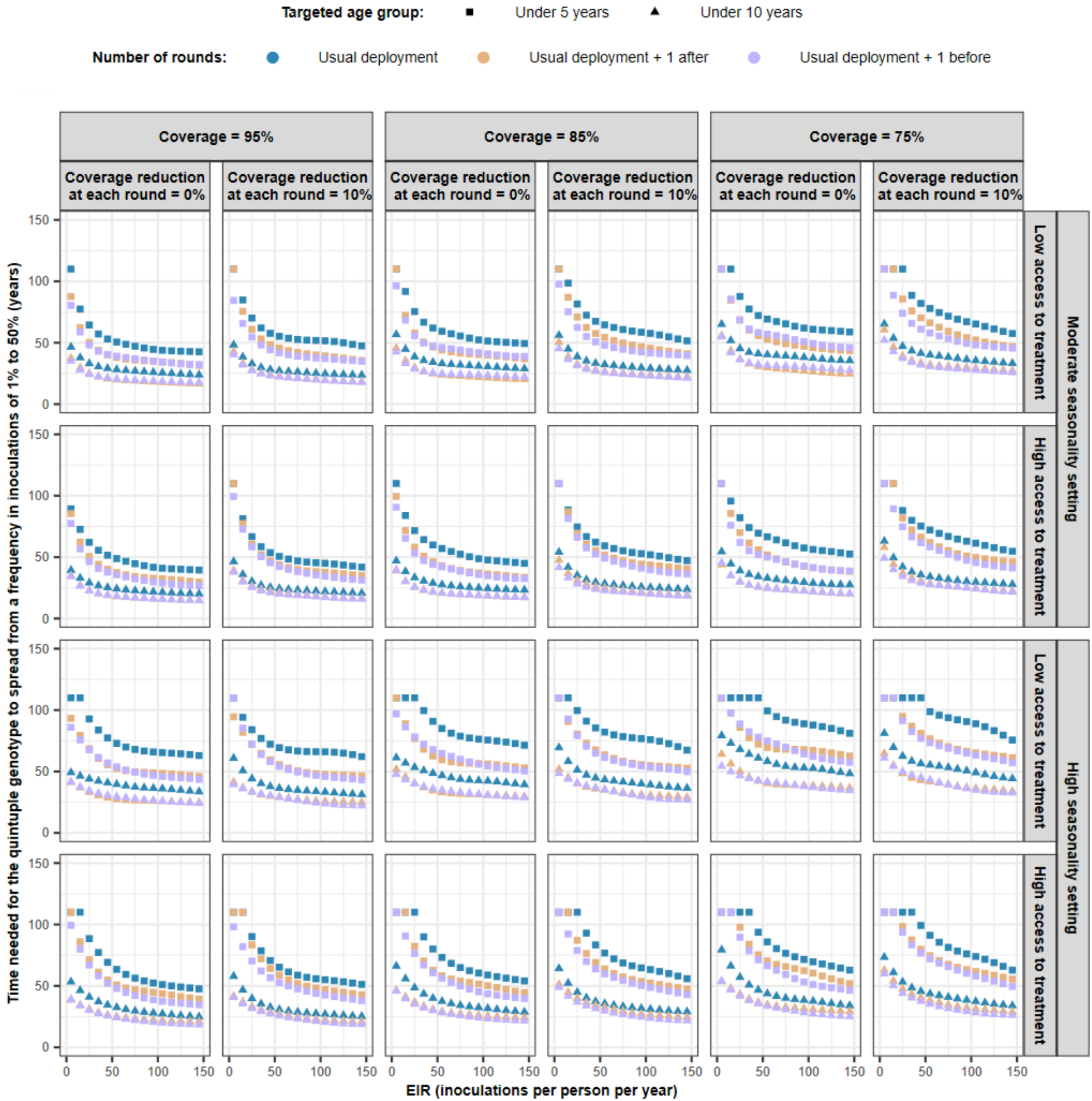
Predicted impact of SMC deployment strategies on spread of the SP-resistant quintuple mutant. Estimated time needed for the quintuple genotype to spread from a frequency in inoculations of 1% to 50%, *T_50_,* when SMC was deployed to different age groups (children aged three months to five years (squares) or children aged three months to ten years (triangles)) in diverse seasonality settings (moderate and high seasonality) with various numbers of cycles deployed per year (high seasonality: three (blue shapes) or four cycles with the additional cycle deployed before (purple shapes) or after (orange shapes) the current deployment period; moderate seasonality setting: four (blue shapes) or five cycles with the additional cycle deployed before (purple shapes) or after (orange shapes) the current SMC deployment period). We predicted *T_50_* for different levels of initial coverage (75%, 85%, and 95%) with coverage reduction across cycles (0% or 10% reduction since the previous cycle). For each setting and SMC deployment strategy, *T_50_* was predicted for various transmission levels (ranging from 5 to 150 inoculations per person per year) for two levels of access to treatment (25% and 70%).

**Figure S2.10:**
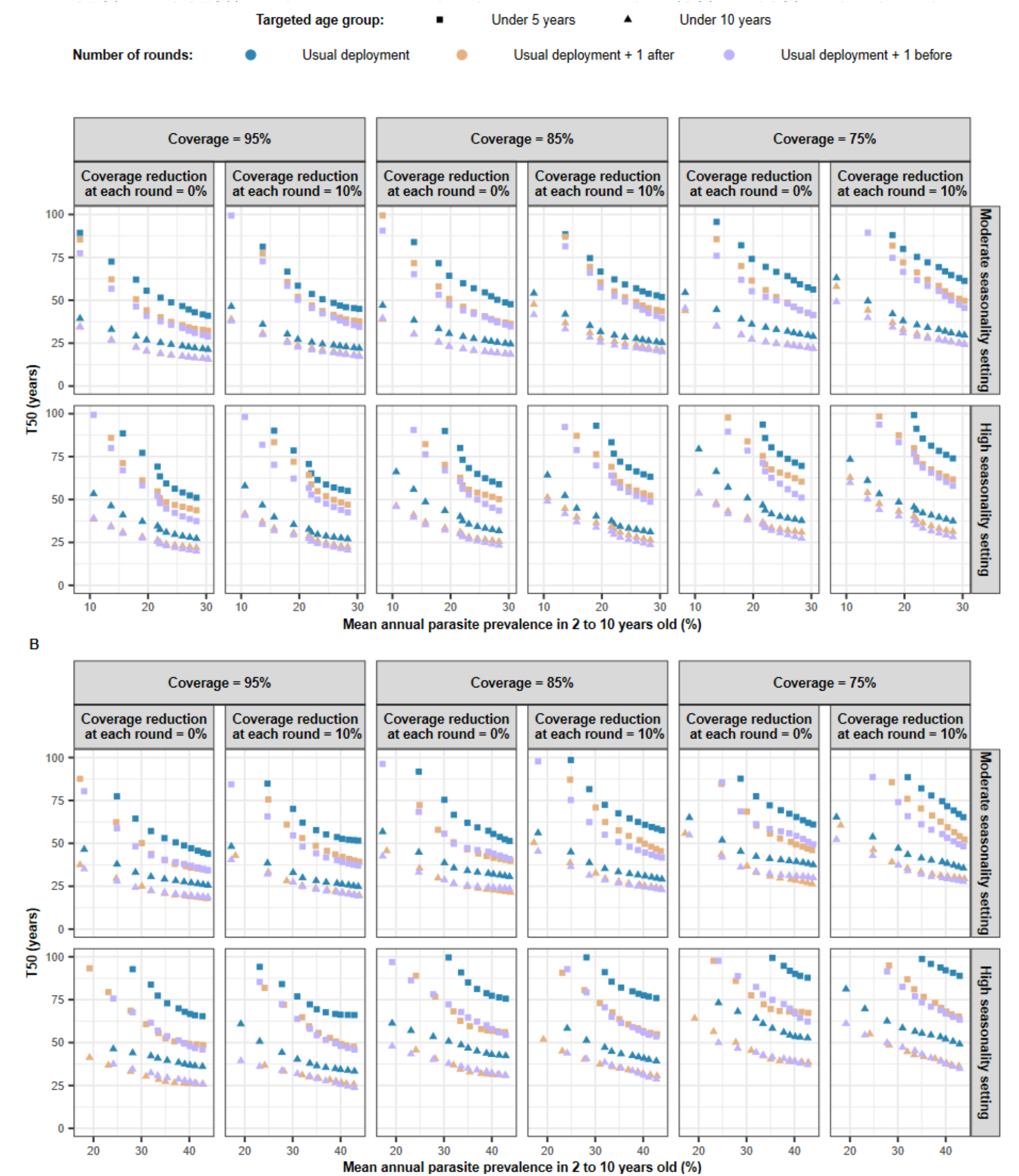
Impact of SMC deployment strategies on the spread of the quintuple mutant. The time needed for the quintuple genotype to spread from a frequency in inoculations of 1% to 50%, *T_50_,* in settings with (A) a high level of access to treatment (70%) and (B) a low level of access to treatment (25%). SMC was deployed to different age groups (children aged three months to five years (squares) or children aged three months to under ten years (triangles)) in diverse seasonality settings (moderate and high seasonality patterns) with a various number of cycles deployed per year (high seasonality: three (blue shapes) or four cycles with the additional cycle deployed before (purple shapes) or after (orange shapes) the standard deployment period; moderate seasonality setting: four (blue shapes) or five cycles with the additional cycle deployed before (purple shapes) or after (orange shapes) the standard SMC deployment period). We predicted the *T_50_* for different levels of initial coverage (75%, 85%, and 95%) and coverage reduction across cycles (0% or 10% reduction since the previous cycle). For each setting and SMC deployment strategy, T50 was predicted for various transmission levels (ranging from 5 to 150 inoculations per person per year) that contributes to variation in prevalence among children between 2 to 10 years old. The mean annual prevalence of P. falciparum was estimated based on the number of patent infections in children between two and ten years.

#### 8 The impact of SMC delivery equity on the spread of the quintuple mutant

In our simulations, we deployed SMC to the same children across cycles, mimicking poor or inequitable SMC delivery, with a proportion of children systematically missed for SMC. We assessed how the *T_50_* changed if SMC was deployed more equitably, that is if all children in the eligible population have the same probability of receiving SMC each cycle within a season. Spread of the quintuple mutant accelerated when SMC was deployed equitably (figure S2.11). This is because, with a more equitable coverage, children reinfected by the quintuple mutant do not necessarily have their infection treated at the next SMC cycle, compared with the scenario with the same children receiving SMC each cycle (inequitable delivery).

**Figure S2.11:**
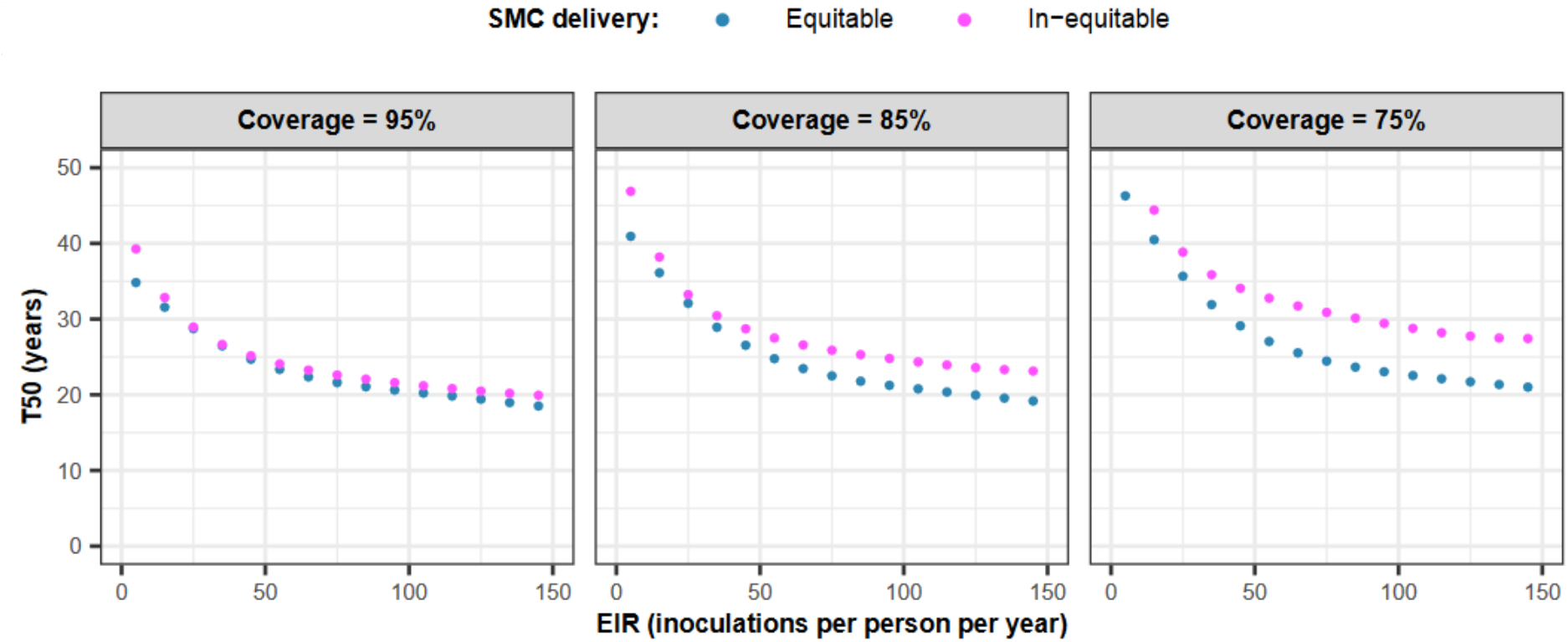
The impact of equitable and inequitable SMC delivery on the spread of the quintuple mutant. We predicted the *T_50_* when delivering SMC with an equitable (blue dots) and an inequitable (pink dots) deployment strategy. We defined an equitable delivery as a deployment where each child had the same probability of receiving SMC each cycle. We defined an inequitable delivery as a deployment where the same children received SMC each cycle. In this illustration, we deployed four cycles of SMC to children aged three months to ten years in a setting with a high access to treatment (70%) and a moderate seasonality pattern of transmission. In each setting and SMC deployment strategy, the *T_50_* was predicted for various transmission levels (ranging from 5 to 150 inoculations per person per year) and three different SMC coverage levels (75%, 85%, and 95%), which were constant across cycles.

#### 9 Estimation of the protective effectiveness of SMC

We estimated the protective effectiveness (PE) of SMC against uncomplicated malaria episodes in parasite populations composed of only one genotype for which SMC provide a prophylactic period of different length (table S2.1). Note that an episode of uncomplicated malaria is the period during which an individual has uncomplicated malaria symptoms caused by parasites present at the time of illness. The maximum length of an episode is 30 days, meaning that illness recurring during this period counts as the same uncomplicated episode.

We simulated the standard deployment of SMC (four cycles of SMC deployed per year to children aged three months to five years in the moderate seasonality setting with a coverage reduction of 10% from the previous cycle) in a human population of 10’000 individuals. Each simulation was run in three seeds, started with a burn-in phase of 130 years, and then we deployed SMC and assessed the PE. To assess the PE, we first estimated the incidence rate (per person per month) of uncomplicated malaria episodes in children between three months and five years during the months of SMC deployment (figure S2.12). For comparison, we estimated the incidence rate of uncomplicated malaria during the same period of the year one year before SMC deployment (figure S2.12). The incidence rates (*I*) were calculated as,

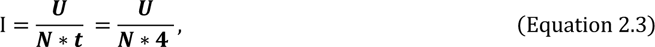

where, *U* is the number of uncomplicated malaria episodes that occur during the four months of SMC deployment, *N* is the number of children aged between three months and five years, and *t* is the number of months of SMC deployment (four). Then, we estimated the PE as the relative reduction of the incidence rate of clinical malaria when SMC was deployed (*I_SMC_*) compared to the incidence rate the year before the deployment (*I_Before SMC_*), as

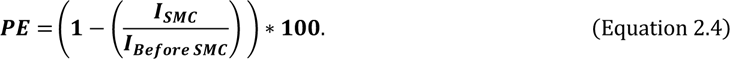

For each prophylactic period length, we assessed the PE for different levels of SMC coverage (75%, 80%, 85%, 90%, 95%, and 100%), access to treatment (10%, 25%, 45%, and 70%), and transmission (ranging between 5 and 150 inoculations per person per year) (figure S2.13).

**Figure S2.12:**
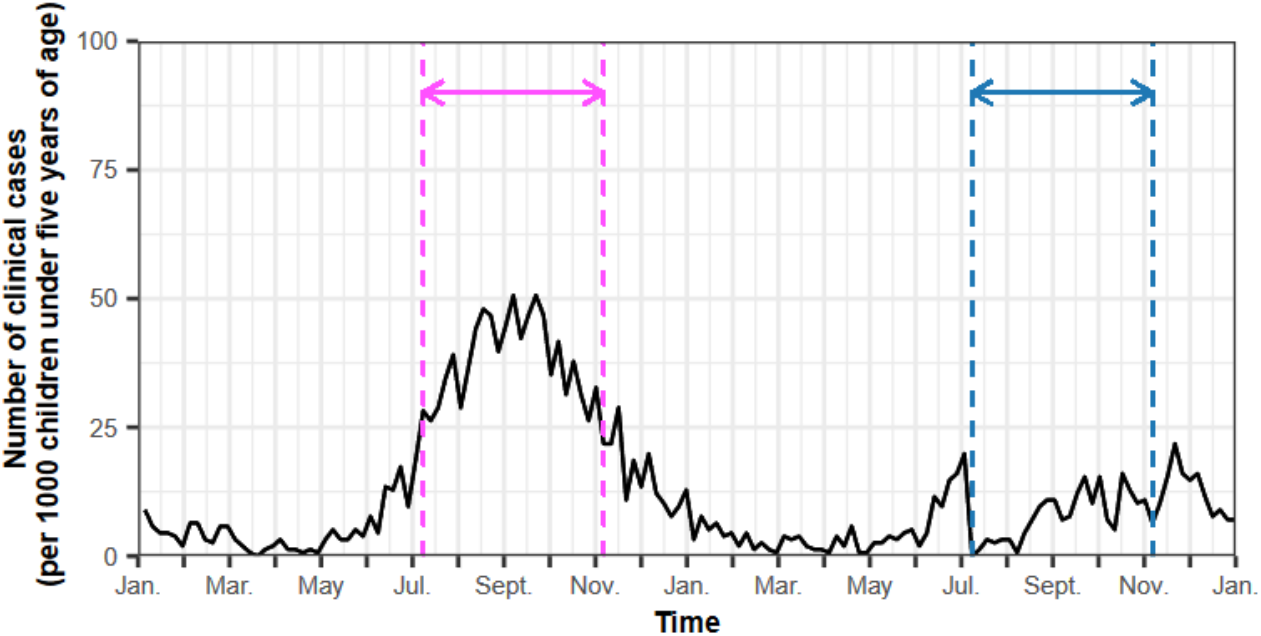
An illustration of the estimation of the SMC protective effectiveness. The black curve represents the incidence of malaria clinical cases per 1000 children aged three months to five years of age over time in the year before the deployment of SMC and the first year of SMC deployment in a parasite population composed only of quadruple mutants (prophylactic period of SP equal to 35 days). The pink arrow and dashed lines indicate the period during which we estimated the incidence rate of clinical malaria before the deployment of SMC. The blue arrow and dashed lines represent the period during which we estimated the incidence of clinical malaria during SMC deployment. In this example, we deployed four cycles of SMC to children aged three months to five years within the period indicated by the blue arrow, with initial coverage of 95%, decreasing by 10% from the previous cycle. The probability of symptomatic cases receiving treatment within two weeks from symptom onset was equal to 25% (low access to treatment). The EIR was equal to 10 inoculations per person per year (medium transmission). The seasonality of the transmission pattern was moderate.

**Table S2.1:** Parameterisation of the EC50 of SP* and AQ to model the desired prophylactic period of SMC. AQ=amodiaquine. DEAQ=desethylamodiaquine. EC50=half-maximal effective concentration. SP*=long acting drug mimicking the synergic action of sulfadoxine-pyrimethamine on the blood-stage.

| Predicted length of the prophylactic period (days) | EC50 of SP* (mg/l) | EC50 of DEAQ (mg/l) | EC50 of AQ (mg/l) |
| --- | --- | --- | --- |
| 5 | 73·450 | 0·130 | 0·0089 |
| 10 | 60·120 | 0·094 | 0·0044 |
| 15 | 42·160 | 0·061 | 0·0044 |
| 21 | 24·200 | 0·024 | 0·0044 |
| 25 | 11·327 | 0·024 | 0·0044 |
| 30 | 4·372 | 0·024 | 0·0044 |
| 35 | 2·39 | 0·024 | 0·0044 |
| 40 | 0·500 | 0·024 | 0·0044 |

blood-stage.

**Figure S2.13:**
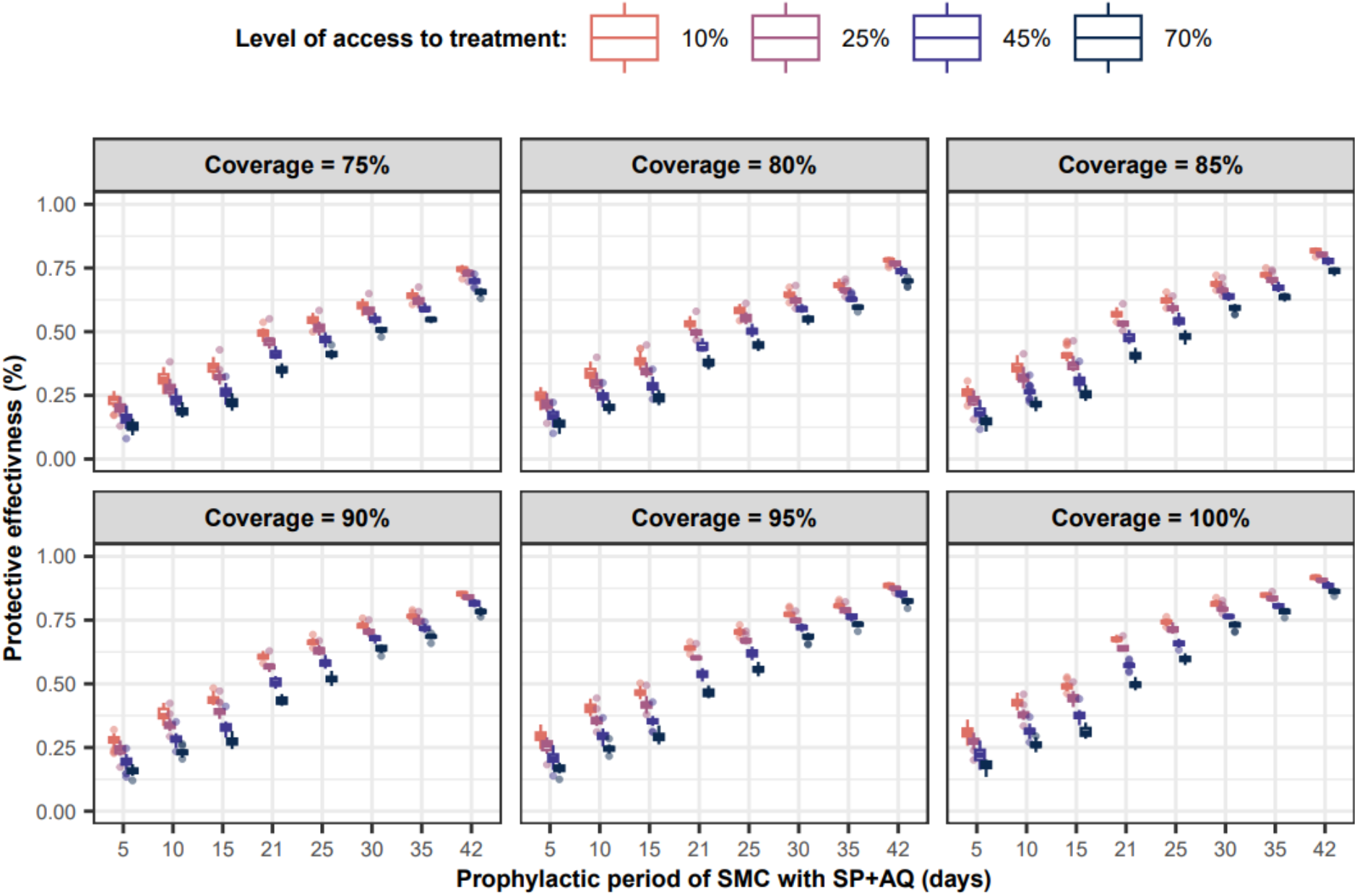
Protective effectiveness of SMC for a range of prophylactic periods. The protective effectiveness of SMC (the relative reduction in the number of clinical malaria cases in children aged three months to five years during the months of SMC implementation) when four cycles of SMC were delivered to these children at different coverage levels (75%, 80%, 85%, 90%, 95%, and 100% with coverage decreased by 10% each cycle), in settings with diverse transmission intensities (from 5 to 150 inoculations per person per year), and at levels of access to treatment from 10% (light blue boxplot) to 70% (dark blue boxplot). The protective effectiveness was assessed against parasite populations composed of 100% of the same genotype for which we varied the PP conferred by SMC with SP+AQ across simulations: 5-, 10-, 15-, 21- (mean PP against quintuple mutants), 25-, 30-, 35- (mean PP against quadruple mutants), and 42- (mean PP against sensitive parasites) days (table S2.1). Note that when SP+AQ provided a PP lower than 17 days, we modelled some degree of resistance to AQ. The x-axis is not equally incremented by five days to illustrate the assumed PP against each genotype. The circles in the boxplots represent outliers.

**Figure S2. 14:**
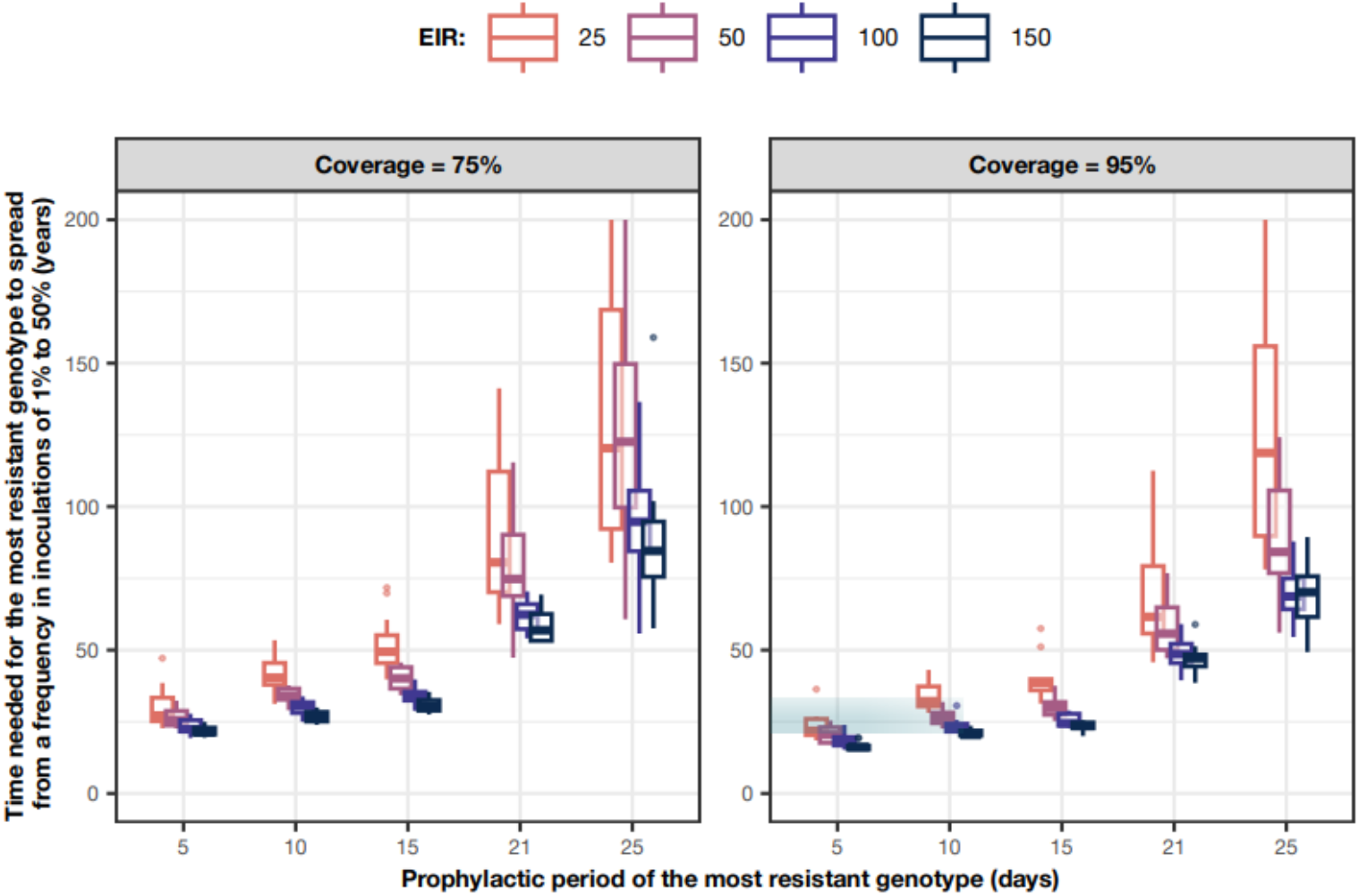
Relationship between the PP of the most resistant genotype and the estimated rate of spread. **We additionally estimated the time needed for resistant genotypes to spread from 1% to 50% frequency of inoculations, *T_50_*,** for various assumptions concerning the PP afforded by SMC against the most resistant genotype: 5, 10, 15, 21 (PP against quintuple mutant assumed in the main analysis of our study) and 25 days. This allowed us to explore our assumptions concerning the PP against the quintuple mutant and to assess how the spread would change if a genotype acquired a higher degree of resistance to SP and some degree of resistance to AQ. In these sensitivity scenarios, if SP+AQ was modelled assuming a PP lower than 17 days, we thus also modelled that the genotype had some degree of resistance to AQ. As with our main analysis, we assumed resistant genotypes spread in a parasite population composed of quadruple mutants for which SP offer a PP of 35 days. The *T_50_* shown in this figure assume four cycles of SMC delivered to children three months to five years at different coverage levels (from 95% to 75% with coverage decreased by 10% from each cycle) in settings with diverse levels of access to treatment (from 10% to 70%), and levels of transmission varying from 25 (orange boxplot) to 150 (dark blue boxplot) inoculations per person per years. The x-axis is not equally spaced in order to illustrate relevant PPs. Circles represent outliers. The blue box and shaded area highlight the example described in the results section.

